# Bounding the average causal effect in Mendelian randomization studies with multiple proposed instruments: An application to prenatal alcohol exposure and attention deficit hyperactivity disorder

**DOI:** 10.1101/2022.05.10.22274902

**Authors:** Elizabeth W Diemer, Alexandra Havdahl, Ole A Andreassen, Marcus R Munafò, Pal R Njolstad, Henning Tiemeier, Luisa Zuccolo, Sonja A Swanson

## Abstract

**Background:** Point estimation in Mendelian randomization (MR), an instrumental variable model, usually requires strong homogeneity assumptions beyond the core instrumental conditions. Bounding, which does not require homogeneity assumptions, is infrequently applied in MR.

**Objective:** We aimed to demonstrate computing nonparametric bounds for the causal risk difference derived from multiple proposed instruments in an MR study where effect heterogeneity is expected,

**Methods:** Using data from the Norwegian Mother, Father, and Child Cohort Study and Avon Longitudinal Study of Parents and Children (n=4457, 6216) to study the average causal effect of maternal pregnancy alcohol use on offspring attention deficit hyperactivity disorder symptoms, we proposed 11 maternal SNPs as instruments. We computed bounds assuming subsets of SNPs were jointly valid instruments, for all combinations of SNPs where the MR model was not falsified.

**Results:** The MR assumptions were violated for all sets with more than 4 SNPs in one cohort and for all sets with more than 2 SNPs in the other. Bounds assuming one SNP was an individually valid instrument barely improved on assumption-free bounds. Bounds tightened as more SNPs were assumed to be jointly valid instruments, and occasionally identified directions of effect, though bounds from different sets varied.

**Conclusions:** Our results suggest that, when proposing multiple instruments, bounds can contextualize plausible magnitudes and directions of effects. Computing bounds over multiple assumption sets underscores the importance of evaluating the assumptions of MR models.

**Synopsis:** *Study question:* Do nonparametric bounds provide useful information in the context of MR studies of prenatal exposures with multiple proposed genetic instruments?

*What’s already known:* Point estimation in MR typically requires strong, unverifiable homogeneity assumptions beyond the core MR assumptions. Bounds, which do not require homogeneity assumptions, are rarely applied in MR.

*What this study adds:* We computed bounds on the average causal effect of alcohol consumption during pregnancy on offspring ADHD symptoms in two European cohorts, proposing 11 genetic variants as instruments. Our results suggest that, when proposing multiple instruments, bounds can contextualize plausible magnitudes and directions of effects.

## Introduction

When estimating causal effects using methods based on confounder adjustment, studies are vulnerable to bias from unmeasured confounding. This is especially problematic for exposure-outcome relationships where confounders are complex or difficult to measure. Mendelian randomization (MR), an instrumental variable model proposing single nucleotide polymorphisms (SNPs) as instruments, is an increasingly popular alternative. Under certain conditions, MR allows for estimation of causal effects even in the presence of unmeasured confounding. Specifically, when proposing a single SNP as an instrument, MR requires that the SNP is associated with the exposure, does not affect the outcome except through the exposure, and individuals at different levels of the SNP are exchangeable with regards to counterfactual outcome ^1^. To obtain a point estimate for the average causal effect in the population, investigators must additionally make one of a set of possible homogeneity assumptions, described in detail elsewhere ^2-5^. Unfortunately, these point estimating conditions are often biologically implausible in MR ^6, 7^.

In contrast, bounding of the average causal effect can be conducted under the 3 primary MR conditions alone. Historically, bounding approaches have been unpopular, possibly because bounds based on a single binary proposed instrument are often wide ^8^. However, when multiple SNPs are proposed as instruments, there are underrecognized opportunities. First, we might tighten bounds by proposing joint sets of SNPs as instruments ^9, 10^. Second, by comparing bounds computed under different assumptions, we might learn more about our reliance on assumptions in informing plausible effect sizes ^11-13^.

This approach may be especially helpful for MR studies of the effect of pregnancy alcohol consumption on offspring outcomes. While several non-MR studies have found positive associations between maternal pregnancy alcohol use and offspring attention deficit hyperactivity disorder (ADHD) ^14-16^, these estimated effects may be confounded by other maternal health behaviors. However, because offspring alcohol exposure depends both on the amount of alcohol consumed by the mother and the speed of the mother’s alcohol metabolism, most versions of homogeneity assumptions required for point estimation using MR are violated when proposing alcohol dehydrogenase-related SNPs as instruments: the effect of alcohol exposure would likely be heterogeneous across offspring of mothers with different genetic variants^6, 8^. Additionally, because the effect of alcohol exposure is likely heterogeneous for other reasons, homogeneity assumptions are also suspect when proposing non-alcohol dehydrogenase SNPs as instruments ^6^. Here, we demonstrate the use of bounds derived from multiple proposed instruments in an MR study where effect heterogeneity is expected, and provide adaptable software for the implementation of the bounds across combinations of proposed instruments.

## Methods

### Data

The Avon Longitudinal Study of Parents and Children (ALSPAC) is a longitudinal birth cohort, which aimed to recruit all pregnant women in former Avon county with a due date between April 1^st^, 1991 and December 31^st^, 1992 and continues to follow the offspring. 75.3% of contacted women agreed to participate, resulting in a total of 14,541 pregnancies enrolled during this period. When the oldest children were approximately 7 years old, the study recruited additional eligible children who had not previously participated. The study now includes data on the offspring of 15,454 pregnancies. Further detail is available elsewhere ^17-19^. The study website contains details on available data through a fully searchable data dictionary and variable search tool (http://www.bristol.ac.uk/alspac/researchers/our-data/). Informed consent for the use of data collected via questionnaires and clinics was obtained from participants following the recommendations of the ALSPAC Ethics and Law Committee at the time. Ethical approval was obtained from the ALSPAC Ethics and Law Committee and the Local Research Ethics Committees. We restricted analyses to singleton pairs of self-reported white European origin with complete data on maternal genotype, maternal pregnancy drinking behavior, and offspring outcomes, resulting in a total analytic sample of 4,457 mother-child pairs (Supplemental Figure 1).

The Norwegian Mother, Father and Child Cohort Study (MoBa) is a population-based pregnancy cohort study conducted by the Norwegian Institute of Public Health. Participants were recruited from all over Norway from 1999-2008. The women consented to participation in 41% of the pregnancies. The cohort now includes 114,500 children, 95,200 mothers, and 75,200 fathers. Detailed information is available elsewhere ^20, 21^. The current study is based on version 12 of the quality-assured data files released for research in January 2019. The establishment of MoBa and initial data collection was based on a license from the Norwegian Data Protection Agency and approval from The Regional Committee for Medical and Health Research Ethics. MoBa is now based on regulations related to the Norwegian Health Registry Act. The current study was approved by The Norwegian Regional Committee for Medical and Health Research Ethics (2016/1702). For this study, we restricted our sample to singleton pairs with complete data on maternal genetics, maternal pregnancy drinking behavior, and offspring outcomes, resulting in a final analytic sample of 6,216 mother-child pairs (Supplemental Figure 2).

### Measures

#### Genetic variants

We selected SNPs based on a recent genome-wide association study of alcohol use in UK Biobank ^22^. Of the 14 SNPs identified at genome-wide significance in that analysis, we excluded 3 SNPs previously found to be associated with traits that could violate MR assumptions via pleiotropy, or were within genes that were associated with such traits ^23-28^. The 11 independent SNPs we thus proposed as instruments were rs145452708, rs193099203, rs11940694, rs29001570, rs3114045, rs140280172, rs9841829, rs35081954, rs9991733, rs149127347, chr18:72124965. ALSPAC mothers were genotyped using the Illumina human660W-quad, and imputed to the 1000 Genome Project. MoBa mothers were genotyped using either Illumina HumanCoreExome or Illumina Global Screening Array, and genotypes were imputed to Haplotype Reference Consortium (HRC) version 1.1. Details of ALSPAC and MoBa genotyping procedures are available in the Supplementary Materials.

In contrast to GWA studies, measurement error of SNPs proposed as instruments will not bias average causal effect estimates of the exposure of interest on the outcome, as long as measurement error of the SNPs is at most differentially associated with the exposure, and not with the outcome ^6^. For this reason, we did not exclude proposed instruments with minor allele frequencies under 5% or imputation quality below 0.8. However, assortative mating can violate MR assumptions ^29^. While Hardy Weinberg equilibrium tests for all SNPs proposed as instruments were conducted as part of the quality control pipeline in both cohorts, these tests may be underpowered to detect small deviations ^30^. However, such deviations could cause large biases in MR. We estimated the correlation between maternal and paternal genotype for each SNP proposed as an instrument in one cohort to identify SNPs which may be particularly vulnerable to this bias (Supplementary Materials).

Because there is incomplete overlap of loci between 1000Genomes and HRC, not all 11 SNPs were available in MoBa. Proxies for unavailable SNPs were selected using LDProxy, based on maximum r^2 31^. Within MoBa, rs145441283 was used as a proxy for rs193099203 and rs1154447 was used as a proxy for rs35081954. Because chr18:72124965 was unavailable in either cohort, rs201288331 was used as proxy in ALSPAC, and rs12955142 was used as a proxy in MoBa.

#### Alcohol Use

Alcohol use in the second and third trimester was assessed via postal questionnaire around gestational weeks 18 and 32 in ALSPAC, in which mothers reported their average volume and frequency of alcohol consumption in the last few weeks. In MoBa, mothers reported average volume and frequency of alcohol consumption between gestational weeks 13-24 and after week 25 via a postal questionnaire at week 30. Although drinking in pregnancy is not truly a binary process, and mild drinking likely incurs different effects than heavy drinking, the bounding approach used here (described below) requires a binary exposure. For that reason, mothers were categorized as ever drinkers if they reported drinking any amount of alcohol during the second or third trimester, and never drinkers if they did not report any drinking during either trimester. Because heavy and moderate drinking were included in the same category, this approach may be vulnerable to bias from poorly defined interventions. To evaluate whether this caused violations of the instrumental inequalities, we applied the instrumental inequalities when grouping alcohol consumption into 3 categories (never drinking, <2 drinks per week, ≥ 2 drinks per week), 4 categories (never drinking, <1 drink per week, 1-2 drinks per week, > 2 drinks per week), and 7 categories (never drinking, <1 drink per week, 1-2 drinks per week, 3-4 drinks per week, 5-6 drinks per week, 7-13 drinks per week, > 13 drinks per week). In secondary analyses, we restricted the study population to compare never drinking and moderate drinking, defined as drinking less than or equal to 32 grams of alcohol per week (approximately 2 drinks per week). Restricting the analytic population in this way can generate selection bias ^32^, which is why this is not the primary approach.

#### ADHD

In ALSPAC, mother-reported ADHD symptoms at age 7 were assessed using the Development and Well-being Assessment ^33^. In MoBa, mother-reported ADHD symptoms at age 5 were assessed using the Child Behavior Checklist attention deficit hyperactivity subscale ^34^. Children with subscale T scores at or above the 98^th^ percentile within the full MoBa cohort (raw score 8, equivalent to the 84^th^ percentile in published norm data) were considered to have ADHD symptoms ^34^. Table 1 shows the prevalence of maternal alcohol use and offspring ADHD symptoms.

**Table 1.** Prevalence of Maternal Alcohol Use and Offspring Attention Deficit-Hyperactivity (ADHD) Symptoms in the Avon Longitudinal Study of Parents and Children (ALSPAC) and the Norwegian Mother, Father, and Child Study (MoBa).

|  | **ALSPAC** | **MoBa** |
| --- | --- | --- |
|  | **% (n)** | **%(n)** |
| n | 4457 | 6216 |
| Alcohol Use During 2<sup>nd</sup> and 3<sup>rd</sup> Trimester of Pregnancy |  |  |
| 0 g/week | 66.9 (1522) | 90.6 (5606) |
| ≤ 32 g/week | 9.5 (216) | 9.0 (555) |
| > 32 g/week | 23.6 (536) | 0.4 (25) |
| Offspring ADHD symptoms | 2.0 (90) | 2.6 (163) |

### Statistical Analysis

When multiple SNPs are believed to be individually valid instruments, several MR models using different subsets of SNPs, and thus slightly different assumptions, are possible. We could conduct MR models separately for each SNP proposed as an instrument. If we were willing to assume several SNPs were individually and jointly valid instruments, we could also conduct MR analyses proposing the set of SNPs as joint instruments. Our analysis plan included computing bounds under combinations of assumptions related to the 10 SNPs being proposed as instruments, as described below. Prior to computing any of these bounds, we applied the instrumental inequalities to attempt to falsify each assumption set^35, 36^. Specifically, in each cohort, we applied the Balke-Pearl instrumental inequalities across all possible combinations of the SNPs proposed as instruments and to a categorical, unweighted allele score, using code developed previously ^7^. We also applied the Bonet instrumental inequalities to each SNP individually. All sets that violated the instrumental inequalities (e.g., resulted in values greater than 1 for the Balke-Pearl inequalities, or greater than 2 for the Bonet inequalities) were eliminated from further analysis. When multiple SNPs are proposed as joint instruments, the MR model can also be falsified if the bounds calculated using the sets flip, meaning the lower bound is higher than the upper bound. Sets that produced flipped bounds were also removed from the results.

As increasingly large numbers of SNPs are proposed as joint instruments, it is increasingly likely that the MR conditions, and thus the instrumental inequalities, will be violated by chance, rather than by a structural bias in the super-population of interest. These random violations are similar to the concept of “random confounding” in randomized control trials^37^. As with random confounding in randomized control trials, if random violations of the MR conditions are present within a sample, an MR analysis in that sample is expected to produce biased effect estimates^7^. By eliminating all sets that violated the instrumental inequalities, we could eliminate all sets for which the MR conditions were clearly falsified. However, because it is unclear which violations of the inequalities represent structural violations of the MR conditions, as opposed to random violations, the extent to which results of the instrumental inequalities in this study can be generalized to other datasets is unclear.

In the setting of a binary exposure and outcome, bounds on the average causal effect can be calculated using exposure and outcome data alone, without any assumptions ^3, 4^. These assumption-free bounds will always have width 1 and always include the null, meaning they cannot identify the direction of effect. Under the MR assumptions, narrower bounds on the average causal effect are possible. When a set of SNPs are assumed to be jointly valid instruments, the set can be combined into a single variable, with levels representing every unique combination of alleles from the included SNPs. This combined variable can then be used to generate bounds using the expression described by Richardson and Robins^10^. To evaluate differences in the bounds across different joint instruments in each cohort, we calculated Richardson-Robins bounds for all combinations of the 11 SNPs that did not violate the instrumental inequalities (Supplementary Materials).

If at least some number k SNPs, but not all 11, were jointly valid instruments, then the average causal effect would lie within the union of the Richardson-Robins bounds computed proposing combinations of k SNPs as joint instruments^9^. To explore this, we computed bounds in each cohort assuming only a subset of the 11 SNPs were jointly valid instruments, for all subset sizes where at least some combinations did not violate the instrumental inequalities.

In the context of alcohol-dehydrogenase related SNPs and prenatal alcohol, the additional homogeneity assumption required for point estimation of the average causal effect in MR is likely invalid. However, in order to explore how conclusions from point estimation and bounding in MR differ, we computed point estimates using two stage least squares (Supplementary Materials).

Although MoBa and ALSPAC are relatively ethnically homogenous, residual population stratification may bias our results. We therefore also calculated the instrumental inequalities and bounds for each possible combination of the proposed instruments using inverse probability weighting to adjust for 10 principal components (Supplementary Materials) ^1^.

All analyses were conducted in R version 3.6.1. Adaptable R code for application of the instrumental variable bounds, filtered by the instrumental inequalities, are available in Supplementary Materials.

## Results

When comparing any alcohol consumption to no alcohol consumption, in ALSPAC, the instrumental inequalities held for all SNPs individually, 28 combinations of 2 SNPs, 16 combinations of 3 SNPs, two combinations of 4 SNPs, and no combinations of 5 or more SNPs (Supplemental Figure 3). In MoBa, the instrumental inequalities held for 9 combinations of 2 SNPs, and did not hold for any combination of 3 or more SNPs (Supplemental Figure 4). In addition, the instrumental inequalities failed to hold for 3 SNPs individually in MoBa. A similar amount and pattern of instrumental inequality violations were observed when comparing moderate alcohol consumption to no alcohol consumption (Supplemental Figures 5-6). Results of the instrumental inequalities were also broadly similar when categorizing alcohol consumption into 3,4, or 7 categories (Supplementary Figures 7-12), and when samples were IP weighted for 10 principal components (Supplementary Figures 13-16).

In ALSPAC, bounds assuming at least one SNP was an individually valid instrument were very wide (−0.52, 0.47), and barely improved on the assumption-free bounds (−0.53,0.47). Bounds calculated using each instrument individually were similarly wide (Figure 1). As the number of SNPs assumed to be jointly valid instruments increased, the bounds narrowed substantially, and sometimes fell completely on one side of the null, identifying the direction of effect. However, bounds from different sets of proposed instruments varied substantially, even identifying opposite directions of effect. With few exceptions, point estimates generally fell within the bounds (Supplementary Table 1).

**Figure 1.**
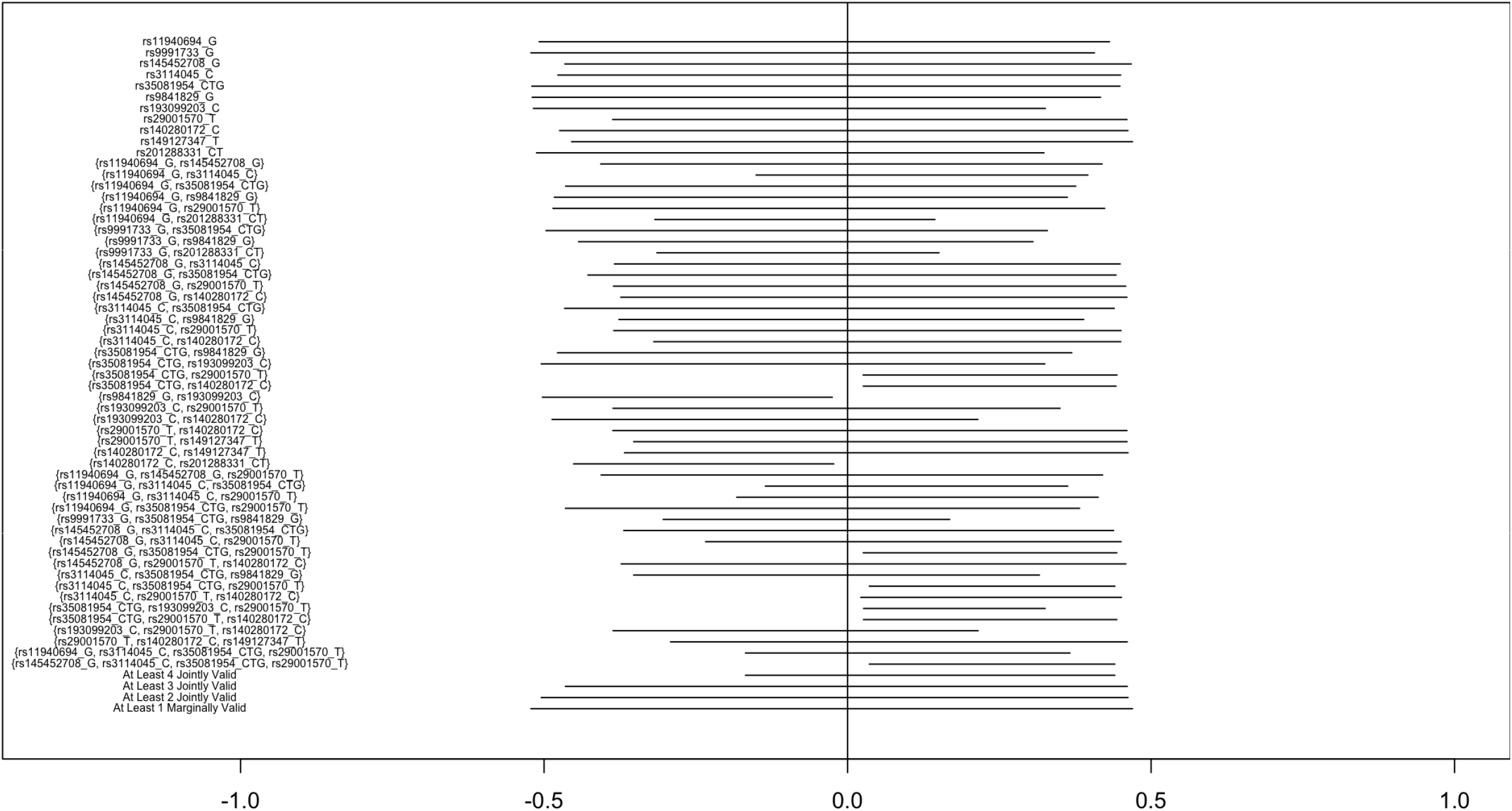
Bounds on the average causal effect of any vs no alcohol consumption during the second and third trimester on offspring attention deficit hyperactivity disorder symptoms in the Avon Longitudinal Study of Parents and Children, proposing different sets of SNPs as instruments.

In MoBa, bounds were consistent across different assumptions (Figure 2). In all cases, the bounds covered the null. In most cases, the bounds did not differ substantially from the assumption-free bounds (−0.12, 0.88), with the narrowest bounds computed being based on the assumption that two SNPs (rs29001570 and rs9841829) were jointly valid (−0.07, 0.73). In 5 of 16 sets of proposed instruments, point estimates fell outside of the bounds (Supplementary Table 2).

**Figure 2.**
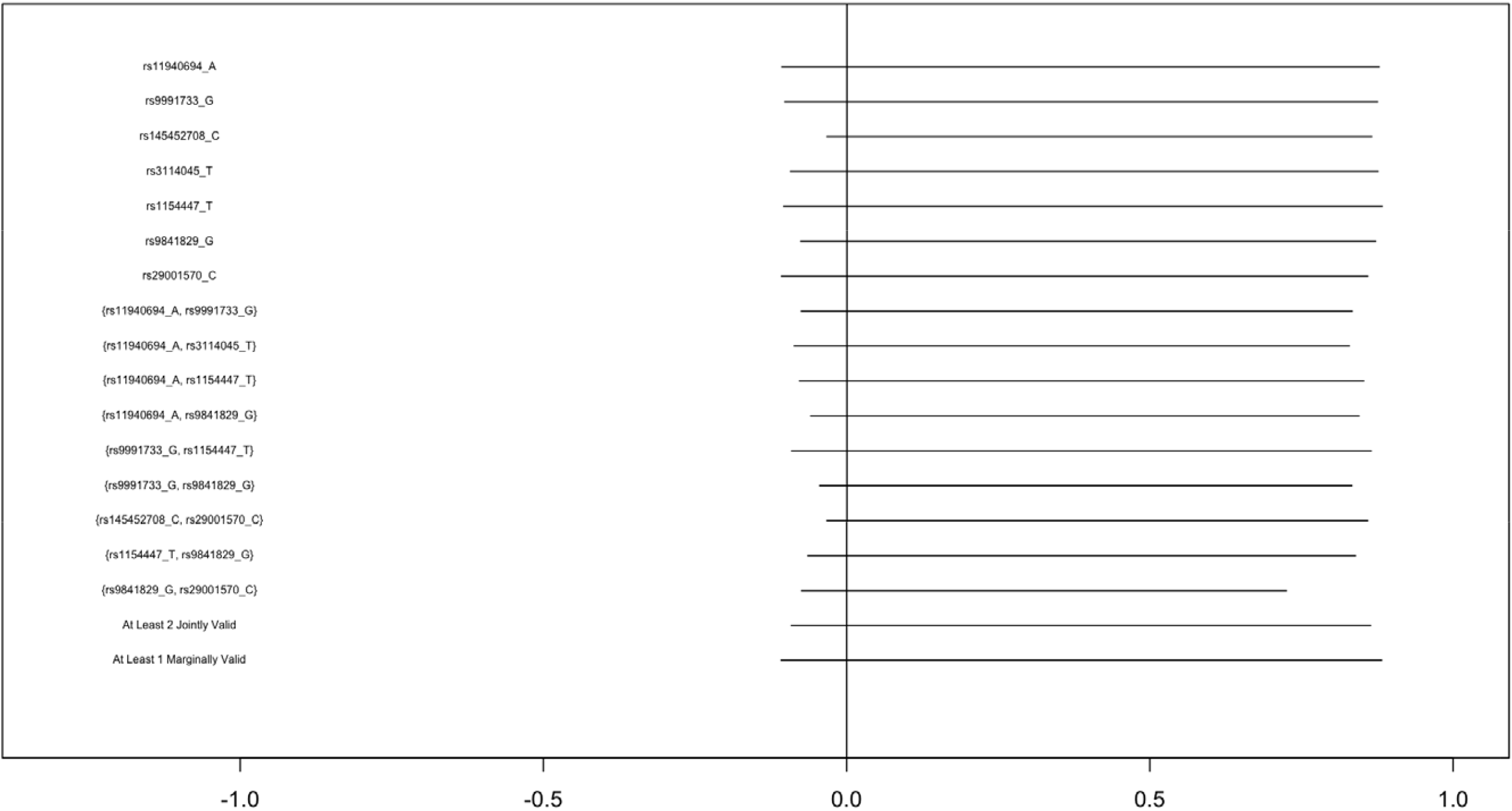
Bounds on the average causal effect of any alcohol vs. no alcohol consumption during the second and third trimesters of pregnancy on offspring attention deficit hyperactivity disorder symptoms in the Norwegian Mother, Father, and Child Study, proposing varying combinations of SNPs as instruments.

Bounds computed to estimate the effect of moderate alcohol consumption, rather than any alcohol consumption, followed a similar pattern in both cohorts (Figures 3 & 4, Supplementary Tables 3-4). In ALSPAC, bounds proposing combinations of 3 or more SNPs narrowed more substantially than the any alcohol models, though bounds still varied substantially and several sets resulted in flipped bounds. Results in each cohort were generally consistent when IP weighted for 10 principal components (Supplementary Figures 17-20, Supplementary Tables 5-8). Correlation between maternal and paternal genotypes was generally very small (Supplementary Table 9).

**Figure 3.**
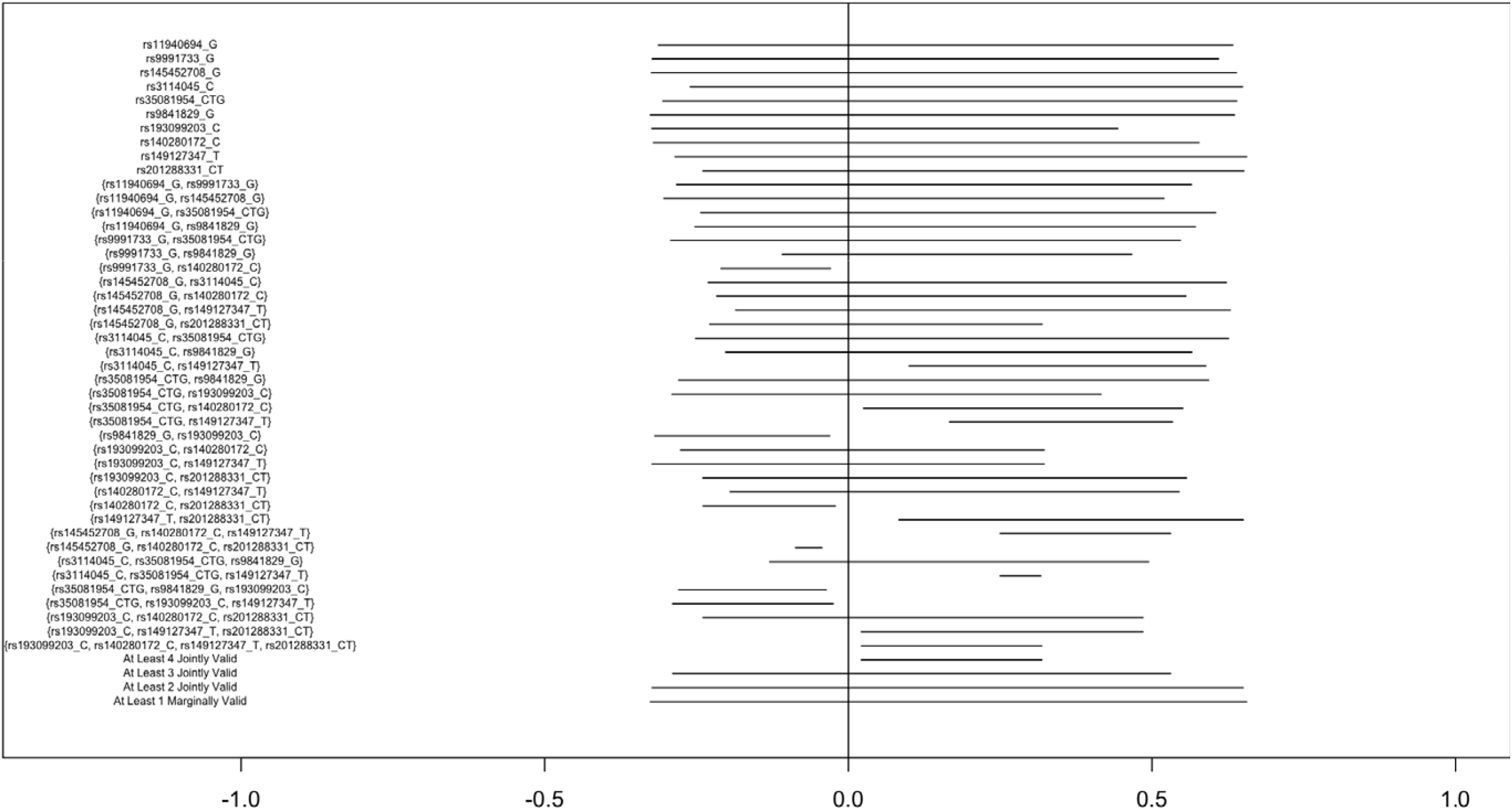
Bounds on the average causal effect of moderate (<2 drinks/week) vs no alcohol consumption during the second and third trimester on offspring attention deficit hyperactivity disorder symptoms in the Avon Longitudinal Study of Parents and Children, proposing varying combinations of SNPs as instruments.

**Figure 4.**
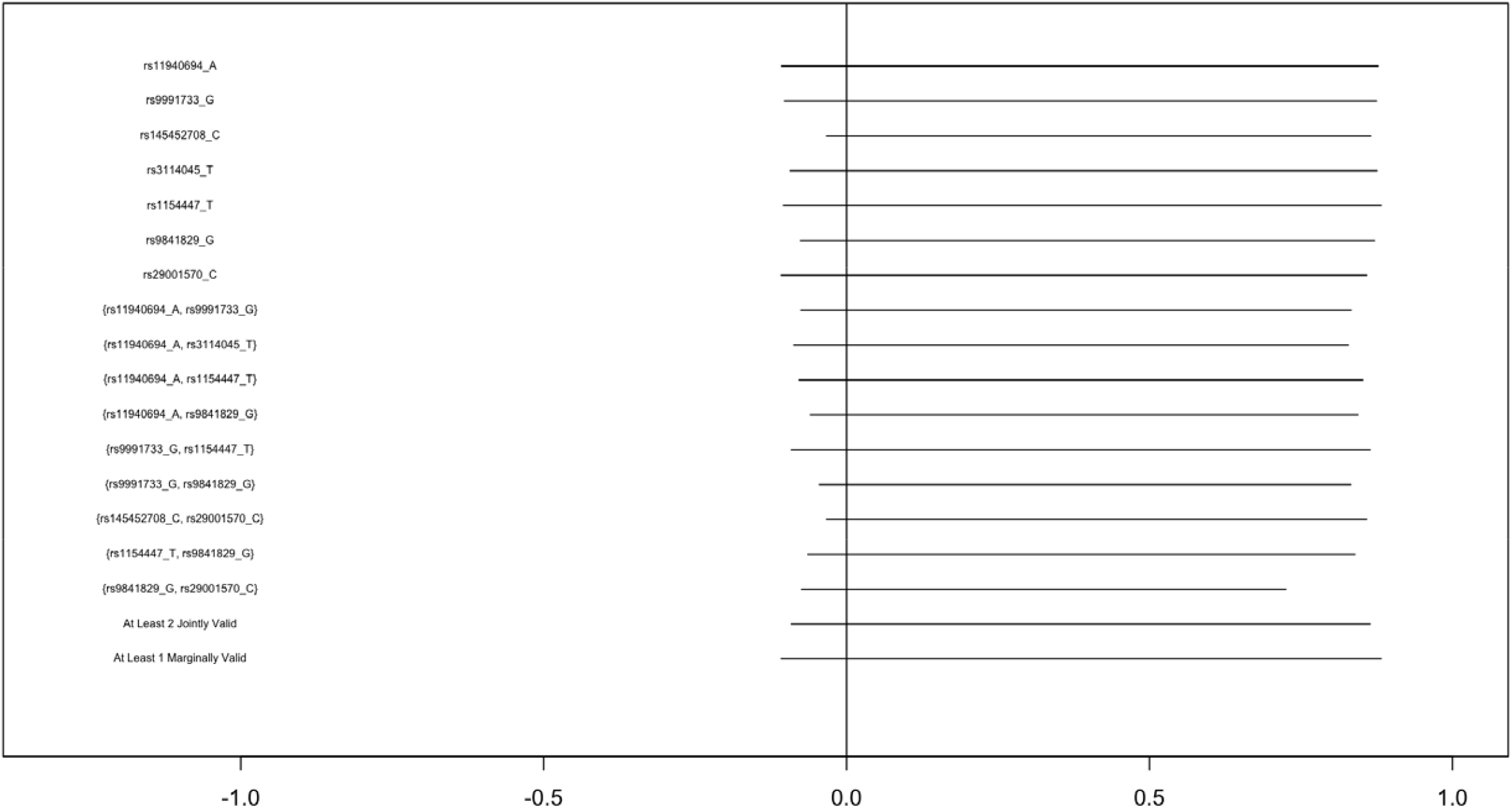
Bounds on the average causal effect of moderate alcohol consumption moderate (<2 drinks/week) vs no alcohol consumption during the second and third trimester on offspring attention deficit hyperactivity disorder symptoms in the Norwegian Mother, Father, and Child Study, proposing varying combinations of SNPs as instruments.

## Discussion

When single SNPs were proposed as instruments, bounds on the average causal effect of both any and moderate prenatal alcohol consumption on offspring ADHD were wide, and were consistent with negative, null, and positive effects. However, in ALSPAC, as increasing number of SNPs were assumed to be joint instruments, bounds narrowed and sometimes identified the direction of effect, though bounds varied substantially across different proposed instruments. In MoBa, the instrumental inequalities held for far fewer sets of proposed instruments compared to ALSPAC. Bounds on the average causal effect of moderate and any alcohol consumption on offspring ADHD remained wide and fairly constant across several different sets of assumptions in MoBa.

Although bounds proposing a single SNP as an instrument barely improved on the assumption-free bounds, the width of the bounds did decrease as we incorporated stronger assumptions. Our ability to evaluate how incorporating stronger assumptions might narrow the bounds was limited by the fact that the strongest assumption sets we considered a priori (that all 11 SNPs were jointly valid instruments) were found to be violated. Nonetheless, bounds in our analysis did narrow as larger numbers of SNPs were proposed as joint instruments, and sometimes identified the direction of effect. This suggests that, when multiple SNPs are proposed as jointly valid instruments, bounds may be able to inform decision-making without additional point estimating assumptions. This may be especially helpful for contexts, like MR studies of prenatal alcohol exposure, where homogeneity assumptions are implausible.

An advantage of computing bounds over many different assumptions is that such approaches can clarify how different assumptions can change study conclusions ^38^. In our application, we were only able to identify a direction of effect under the strong assumption that multiple SNPs were jointly valid instruments. Moreover, in ALSPAC, proposing different sets of SNPs as joint instruments resulted in bounds that identified opposite directions of effects. This variation would have been difficult to identify in many MR point estimation approaches, but is clearly apparent when bounds are evaluated over several possible assumptions. In highlighting these variations, computing bounds over many different assumptions about the SNPs proposed as instruments could shift the focus of MR studies towards the question of what assumptions are most plausible, and thus which range of effects we should be most confident in.

This property may be enhanced by combining bounding with applications of the instrumental inequalities, which could allow for the elimination of analyses based on clearly invalid assumptions ^7^. Our results showed that at least 7 of the SNPs in our analysis could not be valid instruments in ALSPAC, and at least 9 of the 11 could not be valid instruments in MoBa. This is surprising, as the full set of proposed instruments contained 4 SNPs in alcohol dehydrogenase regions, whose relationship to alcohol consumption is relatively well understood. This detected bias could have resulted from several different causes (some of which are detailed in the Supplementary Materials)^39^, but indicates that MR studies of prenatal alcohol exposure may be more vulnerable to bias than was previously understood, and should be viewed with caution. Further investigation is needed to clarify how maternal alcohol-related SNPs impact offspring behavioral health.

The variation in the bounds across assumption sets also illustrates how strongly point estimation in MR relies on the homogeneity assumptions. Even under the strongest unfalsified assumption sets, bounds often covered a moderately large range of effect sizes, meaning point estimation under those sets would still depend heavily on the homogeneity assumptions. Under weaker sets of assumptions, like proposing a single SNP as an instrument, the conclusions of MR studies using point estimation would be informed almost entirely by those additional homogeneity assumptions. This suggests that greater attention should be paid to evaluating the validity of point-estimating assumptions in MR. In our application, point estimates sometimes fell outside the bounds, indicating a violation of the point-identifying assumptions. These sets included SNPs inside and outside of alcohol dehydrogenase regions. While violations of homogeneity were expected in our context, this suggests the resulting bias was severe, and future MR studies might benefit from closer evaluation of the plausibility of the point estimating assumptions.

Even in settings where both the primary MR assumptions and the additional point estimating assumptions are plausible, presentation of the bounds alongside point estimates could help readers and investigators to understand how strongly MR studies depend on assumptions. This is true even, and perhaps especially, when the bounds are wide. Several studies have called for presentation of bounds in observational studies, particularly for instrumental variable models like MR ^11-13, 38^. Robins and Greenland noted that “wide bounds make clear the degree to which public health decisions are dependent on merging the data with strong prior beliefs” ^12^. Incorporating bounds into MR practice would clarify the amount of information present in the data alone, and the need for critical evaluation of assumptions within each study’s unique context.

Further research is needed to extend bounding approaches for instrumental variables and MR in several ways, including but not limited to extensions for: estimation procedures incorporating sampling variability^11, 40^; time-varying interventions^41-43^; conditional instrumental variables incorporating measured covariates^6^; non-binary exposures^44, 45^; and two-sample approaches^46^. Though this list is not exhaustive, we believe it represents priorities for maximizing the usefulness and applicability of bounding in MR.

### Conclusion

Our results show that, when multiple SNPs are proposed as instruments, it is possible to narrow bounds on the average causal effect. The extent of this narrowing will likely depend on the study question and population, but sometimes may allow for identification of the direction of effect. Further, the variation of the bounds observed across different proposed instruments provides a clear example of how bounding can be used to evaluate how heavily an MR analysis depends on assumptions regarding a particular SNP.

MR studies frequently propose large numbers of SNPs as joint instruments, and thus make equivalently large numbers of assumptions about the joint validity of those proposed instruments. Adding to the growing arsenal of sensitivity analyses, bounding may allow researchers to leverage these assumptions to make meaningful conclusions about effects without additional homogeneity assumptions. Even when homogeneity assumptions are biologically plausible, estimating bounds across different combinations of proposed instruments may allow investigators to better evaluate the dependence of their conclusions on those assumptions.

## Data Availability

ALSPAC data access is through a system of managed open access. The following steps highlight how to apply for access to the data included in this data note and all other ALSPAC data:
1. Please read the ALSPAC access policy (http://www.bristol.ac.uk/media-library/sites/alspac/documents/researchers/data-access/ALSPAC_Access_Policy.pdf) which describes the process of accessing the data and samples in detail, and outlines the costs associated with doing so.
2. You may also find it useful to browse our fully searchable research proposals database (https://proposals.epi.bristol.ac.uk/?q=proposalSummaries), which lists all research projects that have been approved since April 2011.
3. Please submit your research proposal (https://proposals.epi.bristol.ac.uk/) for consideration by the ALSPAC Executive Committee.
MoBa data are used by researchers and research groups at both the Norwegian Institute of Public Health and other research institutions nationally and internationally. The research must adhere to the aims of MoBa and the participants given consent. All use of data and biological material from MoBa is subject to Norwegian legislation. More information can be found on the study website (https://www.fhi.no/en/studies/moba/for-forskere-artikler/research-and-data-access/).

http://www.bristol.ac.uk/media-library/sites/alspac/documents/researchers/data-access/ALSPAC_Access_Policy.pdf

## Acknowledgements

We thank Miguel Hernan for his helpful feedback on earlier drafts of this work.

We thank the Norwegian Institute of Public Health (NIPH) for generating high-quality genomic data. This research is part of the HARVEST collaboration, supported by the Research Council of Norway (NRC) (#229624). We also thank the NORMENT Centre for providing genotype data, funded by NRC (#223273), South East Norway Health Authority and KG Jebsen Stiftelsen. Further we thank the Center for Diabetes Research, the University of Bergen for providing genotype data and performing quality control and imputation of the data funded by the ERC AdG project SELECTionPREDISPOSED, Stiftelsen Kristian Gerhard Jebsen, Trond Mohn Foundation, NRC, the Novo Nordisk Foundation, the University of Bergen, and the Western Norway health Authorities (Helse Vest). The Norwegian Mother and Child Cohort Study is supported by the Norwegian Ministry of Health and Care Services and the Ministry of Education and Research. We are grateful to all the participating families in Norway who take part in this on-going cohort study. This work was performed on the TSD (Tjeneste for Sensitive Data) facilities, owned by the University of Oslo, operated and developed by the TSD service group at the University of Oslo, IT-Department (USIT).

We are extremely grateful to all the families who took part in the ALSPAC study, the midwives for their help in recruiting them, and the whole ALSPAC team, which includes interviewers, computer and laboratory technicians, clerical workers, research scientists, volunteers, managers, receptionists and nurses.

