## Supplementary Materials for "Bounding the average causal effect in Mendelian randomization studies with multiple proposed instruments: An application to prenatal alcohol exposure and attention deficit hyperactivity disorder"

#### Contents

1 **Additional assumptions for point estimation in MR** **1**

2 **Description of genotyping in ALSPAC** **2**

3 **Description of genotyping in MoBa** **2**

4 **Expression for Richardson-Robins bounds for all possible combinations of instruments** **3**

5 **Point estimation procedures** **4**

6 **Expression of inverse probability weights for each proposed joint instrument** **4**

7 **Possible violations of the MR assumptions in this analysis** **4**

8 **Supplementary figures 1-20** **6**

9 **Supplementary tables 1-9** **19**

10 **R functions for application of bounds across multiple proposed instruments** **31**

**References** **48**

### 1 Additional assumptions for point estimation in MR

In order to obtain a point estimate using MR, researchers make one of a set of additional homogeneity assumptions. The details of these assumptions, and their relative advantages and disadvantages, have been described in detail elsewhere (Hernan and Robins 2020; Swanson et al. 2018; Wang and Tchetgen 2018). In brief, researchers assume one of the following:

4a. A constant effect of $X$ on $Y$ across individuals
4b. $E(Y^{x=1} - Y^{x=0}|Z = 1, X = x) = E(Y^{x=1} - Y^{x=0}|Z = 0, X = x)$ for all x.

4c. $E(Y^{x=1}|X = x, Z = 1)/E(Y^{x=0}|X = x, Z = 1) = E(Y^{x=1}|X = x, Z = 0)/E(Y^{x=1}|X = 0, Z = 0)$

4d. $E(Y^{x=1}|U) - E(Y^{x=0}|U) = E(Y^{x=1}) - E(Y^{x=0})$

4e. $E(X|Z = 1, U) - E(X|Z = 0, U) = E(X|Z = 1) - E(X|Z = 0)$

If these assumptions are unreasonable, researchers could instead assume monotonicity:

4f. $X^z$ is a nondecreasing function of $z$ on the support of $Z$.

However, under 4f, researchers are only able to identify the average causal effect within the compliers, those individuals for whom $X^{z=a} > X^{z=b}$ for all $a > b$.

Importantly, with the exception of 4c, the choice of assumption will not alter the usual IV estimand, but only the interpretation of the estimate.

As discussed in the main text, 4b (and 4c) are unlikely to hold in the context of alcohol metabolism-related genetic variants proposed as instruments. Beyond this, it is important to recognize that 4a-4f are all strong and unverifiable assumptions, which should be applied with caution. Nonetheless, where there is evidence that the homogeneity assumptions may be reasonable, researchers might consider presenting bounds alongside point estimates computed under assumptions of additive and multiplicative effect homogeneity, in order to see the fullest possible picture of the potential impact of different forms of heterogeneity on their results.

### 2 Description of genotyping in ALSPAC

ALSPAC children were genotyped using the Illumina HumanHap550 quad chip genotyping platforms. The resulting raw genome-wide data were subjected to standard quality control methods. Individuals were excluded on the basis of gender mismatches; minimal or excessive heterozygosity; disproportionate levels of individual missingness (>3%) and insufficient sample replication (IBD < 0.8). Population stratification was assessed by multidimensional scaling analysis and compared with Hapmap II (release 22) European descent (CEU), Han Chinese, Japanese and Yoruba reference populations; all individuals with non-European ancestry were removed. SNPs with a call rate of < 95% or evidence for violations of Hardy-Weinberg equilibrium (P < 5E-7) were removed. Cryptic relatedness was measured as proportion of identity by descent (IBD > 0.1). Related subjects that passed all other quality control thresholds were retained during subsequent phasing and imputation. 9,115 subjects and 500,527 SNPs passed these quality control filters.

ALSPAC mothers were genotyped using the Illumina human660W-quad array at Centre National de Génotypage (CNG) and genotypes were called with Illumina GenomeStudio. PLINK (v1.07) was used to carry out quality control measures on an initial set of 10,015 subjects and 557,124 directly genotyped SNPs. SNPs were removed if they displayed more than 5% missingness or a Hardy-Weinberg equilibrium P value of less than 1 x $10^{-06}$. Samples were excluded if they displayed more than 5% missingness, had indeterminate X chromosome heterozygosity or extreme autosomal heterozygosity. Samples showing evidence of population stratification were identified by multidimensional scaling of genome-wide identity by state pairwise distances using the four HapMap populations as a reference, and then excluded. Cryptic relatedness was assessed using a IBD estimate of more than 0.125 which is expected to correspond to roughly 12.5% alleles shared IBD or a relatedness at the first cousin level. Related subjects that passed all other quality control thresholds were retained during subsequent phasing and imputation. 9,048 subjects and 526,688 SNPs passed these quality control filters.
After combining genotype data in the mothers and the children, SNPs with genotype missingness above 1% were removed due to poor quality (11,396 SNPs removed) and a further 321 subjects were removed due to potential ID mismatches. This resulted in a dataset of 17,842 subjects. Imputation of the target data was performed using Impute V2.2.2 against the 1000 genomes reference panel (Phase 1, Version 3) (all polymorphic SNPs excluding singletons), using all 2186 reference haplotypes (including non-Europeans).
This gave 8,196 eligible mothers with available genotype data after exclusion of related subjects using cryptic relatedness measures described previously.

### 3 Description of genotyping in MoBa

Genotyping of MoBa participants is currently ongoing, and this analysis was conducted using the first available maternal genetic data. Approximately 17,000 trios from MoBa were genotyped in 3 batches. Samples were selected randomly, and excluded from genotyping if the trio met any of the following exclusion criteria: 1) offspring stillborn, 2) offspring deceased, 3) twin offspring, 4) non-existent Medical Birth Registry data, 5) missing anthropometric measures at birth in Medical Birth Registry, 6) pregnancies where the mother did not answer the first questionnaire (as a proxy for higher fallout rate), 7) missing parental DNA samples. The first batch, comprising 20,664 individuals (including parents and children), was genotyped at the Genomics Core Facility (Iceland) using the Illumina HumanCoreExome (Illumina, San Diego, USA) genotyping array, version 12 1.1. The second batch, comprising 12,874 individuals, was genotyped at the Genomics Core Facility (Iceland) using the Illumina HumanCoreExome (Illumina, San Diego, USA) genotyping array, version 24 1.0. The third batch, comprising 17,949 individuals, was genotyped at Erasmus MC (the Netherlands) using the Illumina Global Screening Array (Illumina, San Diego, USA) version 24 1. Genotypes were called using GenomeStudio (Illumina, San Diego, USA) and converted to PLINK format files.

PLINK version 1.90 beta 3.36 (http://pngu.mgh.harvard.edu/purcell/plink/) was used to conduct the quality control, which has been previous described by Helgeland et al (Helgeland et al. 2019). Known problematic SNPs previously reported by the Cohorts for Heart and Aging Research in Genomic Epidemiology consortium and Psychiatric Genomics Consortium were excluded from each batch. Duplicate samples were removed, and each batch was split into parents and offspring. Quality control was conducted separately for parents and offspring.

Individuals were excluded if they had a genotyping call rate below 955 or autosomal zygosity greater than four standard deviations from the sample mean. SNPs were excluded if they were ambiguous, had a genotyping call rate below 98%, or Hardy-Weinberg equilibrium p-value less than 1 x $10^{-6}$. Population stratification was assessed using the HapMap phase 3 release 3 as a reference, by principal component analysis using EIGENSTRAT version 6.1.4. Visual inspection identified a homogenous population of European ethnicity, and individuals of non-european ethnicity were removed. A sex check was done by assessing the sex declared in the pedigree with the genetic sex, which was imputed based on the heterozygosity of chromosome X. When sex discrepancies were identified, the individual was flagged. Relatedness was assessed by flagging one individual from each pairwise comparison of identity-by-descent with a pi-hat greater than 0.1.

Parent and offspring datasets were then merged into one dataset per genotyping batch. All individuals passing the genotyping call rate and normal heterozygosity measures were included in the merged datasets, meaning individuals who had previously been flagged or excluded for being a duplicate, having a sex discrepancy, being an ethnic outlier, or having a high level of relatedness, were included. Concordance checks were then conducted on validated duplicates. Duplicate and tri-allelic SNPs, as well as SNPs that were discordant between validated duplicates were excluded. Indivudals with a genotyping call rate below 98% in the merged datasets were removed. Insertions and deletions were excluded.

Phasing was conducted using Shapeit 2 release 837 and the duoHMM approach was used to account for pedigree structure. Imputation was conducted using the Haplotype Reference Consortium release 1.1 as the reference panel. The Sanger Imputation Server was used to perform the imputations with the Positional Burrows-Wheeler Transform. Phasing and imputation were conducted separately for each genotyping batch.

Imputation quality control was performed by initially converting dosages to best-guess genotypes. Individuals were removed if they had a genotyping call rate less than 99% or were of non-European ethnicity. SNPs with a genotyping call rate less than 98%, or a Hardy-Weinberg equilibrium p-value less than 1 x $10^{-6}$ were removed. Relatedness was assessed intergenerationally and across batches by flagging one individual from each pairwise comparison of identity-by-descent with a pi-hat greater than 0.15 (excepting known parent-offspring relationships). Individuals were flagged for removal only if the other member of the pair would otherwise be included in the same analysis. One individual from each pair was flagged at random, except when retaining one individual would keep more duo/trio data available, in which case the other member was dropped. After quality control, a core homogenous sample of European ethnicity (based on PCA of markers overlapping with available HapMap markers) individuals across all batches and array were available for analysis, resulting in a total n of 14,804 mothers prior to analysis-specific exclusions.

### 4 Expression for Richardson-Robins bounds for all possible combinations of instruments

Richardson and Robins (2014) considered a model in which $X$ and $Y$ are binary, taking states $\{0,1\}$, and $Z$ takes states $\{1,2,..,k\}$ under 4 different assumptions:
(i) $Z \perp\!\!\!\perp Y^{x_1}, Y^{x_0}, X^{z_1}, ..., X^{z_k}$
(ii) $Z \perp\!\!\!\perp Y^{x_0}, Y^{x_1}$
(iii) for $i \in \{1,...,k\}, j \in \{0,1\}, Z \perp\!\!\!\perp X^{z_i}, Y^{x_j}$
(iv) there exists a $U$ such that $U \perp\!\!\!\perp Z$ and for $j \in \{0,1\}, Y^{x_j} \perp\!\!\!\perp X, Z|U$

Each of these assumptions is a slightly different version of the IV conditions used in the literature (Swanson et al. 2018). Under assumption (i), (ii), (iii), or (iv), for all $i,j \in 0,1$, $P(Y^{x_i} = j) \leq g(i,j)$ where

$g(i,j) = min\{min_z[P(X=i, Y=j|Z=z) + P(X=1-i, |Z=z)],$

$\min_{z,\tilde{z}:z\neq\tilde{z}}[P(X=i,Y=j|Z=z)+P(X=1-i,Y=0|Z=z)+P(X=i,Y=j|Z=\tilde{z})+P(X=1-i,Y=1|Z=\tilde{z})]\}$

Because $P(Y^{x_0})$ and $P(Y^{x_1})$ are variation independent, the average causal effect of X on Y, denoted $ACE(X \rightarrow Y)$, is bounded by

$1-g(1,0)-g(0,1) \leq ACE(X \rightarrow Y) \leq g(0,0)+g(1,1)-1$

Returning to our setting with multiple proposed instruments, we can consider the set of proposed instruments $B=\{b_1,b_2,..,b_n\}$. We note that any combination of the proposed instruments in $B$ that are themselves categorical variables can be combined into a single joint instrument $Z$ which takes states $\{1,2,..,k\}$, where each state is a unique possible combination of values of the proposed joint instruments in the subset. Thus the Richardson-Robins bounds can be applied to any joint instrument $Z$, assuming (i), (ii), (iii), or (iv) hold both individually and jointly for each proposed instrument included in $Z$. In our application, we considered this for all possible subsets of our set of proposed instruments.

### 5 Point estimation procedures

For each proposed joint instrument $Z_l$, point estimates for the average causal effect were estimated using two-stage least squares, using linear regression models for both steps. 95% confidence intervals were estimated using basic bootstrap. Two stage squares were estimated using the ivreg() function from the AER package (Kleiber, Zeileis, and Zeileis 2020), and bootstrapping was conducting using the boot.ci() function from the boot package (Ripley 2010).

In the context of categorical exposures and outcomes, two stage least squares using linear regression is vulnerable to measurement error, which can result in predicted values of the exposure outside of the 0-1 range, and may violate the assumption of bivariate normally distributed errors (Rassen et al. 2009). However, some research has suggested this issue may be primarily theoretical, and has limited impact on practical applications (Rassen et al. 2009; Angrist 2001; Johnston et al. 2008) . Some MR researchers attempt to avoid this issue by using models based on logistic regression. However, these approaches will produce estimates of the causal odds ratio, rather than the average causal effect on the risk difference scale. In order to produce point estimates of the average causal effect on the same risk difference scale as the bounds, we therefore chose to use two stage least squares based on linear regression. The resulting point estimates are shown in Supplementary Tables 1-4.

### 6 Expression of inverse probability weights for each proposed joint instrument

For each proposed joint instrument $Z_l$ , unstabilized inverse probability weights (Robins 1997) to account for 10 principal components were estimated as follows:

$W^A = 1/P(Z_l|PC_1,PC_2,PC_3,PC_4,PC_5,PC_6,PC_7,PC_8,PC_9,PC_{10})$

To estimate $W^A$, we fitted multinomial logistic regression models predicting $Z_l$ assuming the principal components contributed additively and linearly on the logit scale. Values were subsequently back-transformed to probabilities, and we calculated
$1/P(Z_l|PC_1,PC_2,PC_3,PC_4,PC_5,PC_6,PC_7,PC_8,PC_9,PC_{10})$ for each individual using these back-transformed probabilities.

### 7 Possible violations of the MR assumptions in this analysis

The MR assumptions are strong, and any or all the SNPs proposed as instruments in our analysis may have been affected by a number of different biases. While the complete case approach used in this analysis aligns with common practice in MR, both analytic samples were much smaller than the original recruited cohort, meaning the results may have been affected by selection bias due to loss to followup and missing data.

Further, because both cohorts were recruited based on the presence of a pregnancy, and offspring ADHD status can only be evaluated in women who become pregnant and carry to term, the MR conditions would have been violated if a woman's alcohol consumption impacted her probability of becoming pregnant (Diemer et al. 2020). Results were largely consistent when inverse probability weighted for 10 principal components, suggesting that the results were not affected by residual population stratification. The correlation between maternal and paternal genotype was small, implying the study was not strongly biased by assortative mating. While the MR assumptions can be violated if offspring outcomes are affected by offspring genotype, this path was unlikely to have impacted our analyses, because alcohol dehydrogenase genes are not expressed in fetuses or young children, who process alcohol through a different mechanism (Faassen and Niemelä 2011). The MR conditions could also be violated if maternal genetic variants impacted offspring ADHD through mechanisms other than alcohol consumption, such a consumption of other substances, if maternal alcohol consumption after birth also impacted offspring ADHD, or if the relationship between maternal genotype and alcohol consumption changed over the course of pregnancy. As mentioned in the main text, if the relationship between maternal alcohol use and offspring ADHD was truly continuous, binarizing alcohol use could violate the MR conditions. However, results of the instrumental inequalities were consistent across increasing numbers of categories of alcohol use, suggesting violations of the instrumental inequalities were not primarily due to our categorization of alcohol use. While it is possible that the MR conditions could also be violated as a result of measurement error of exposure, maternal alcohol use, previous research suggests bias from such measurement error is generally mild (Pierce and VanderWeele 2012).

#### 8 Supplementary figures 1-20

Supplementary figure 1:
Flowchart of Avon Longitudinal Study of Parents and Children included in analytic sample

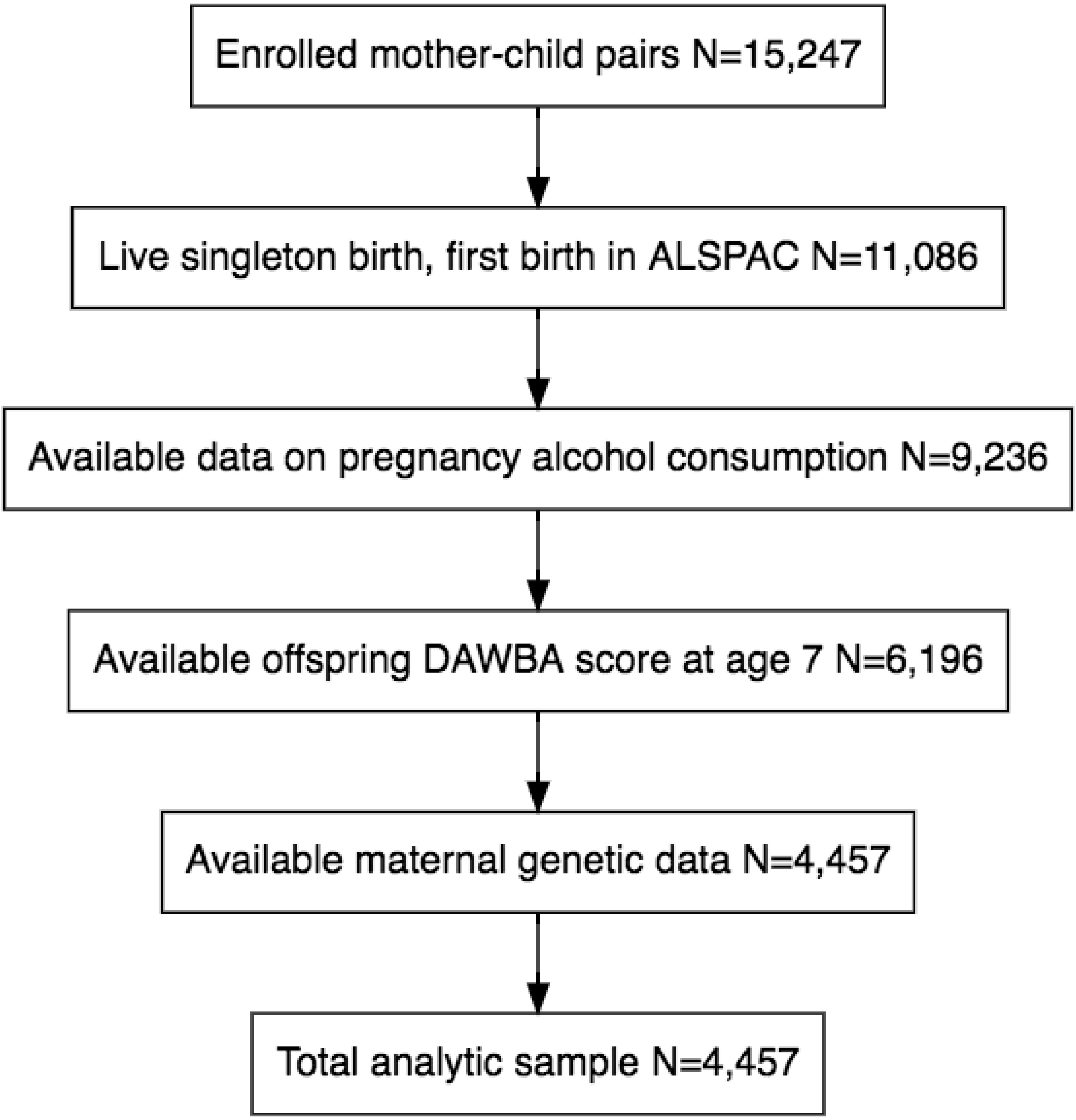

Supplementary figure 2: Flowchart of Norwegian Mother, Father, and Child Study included in analytic sample

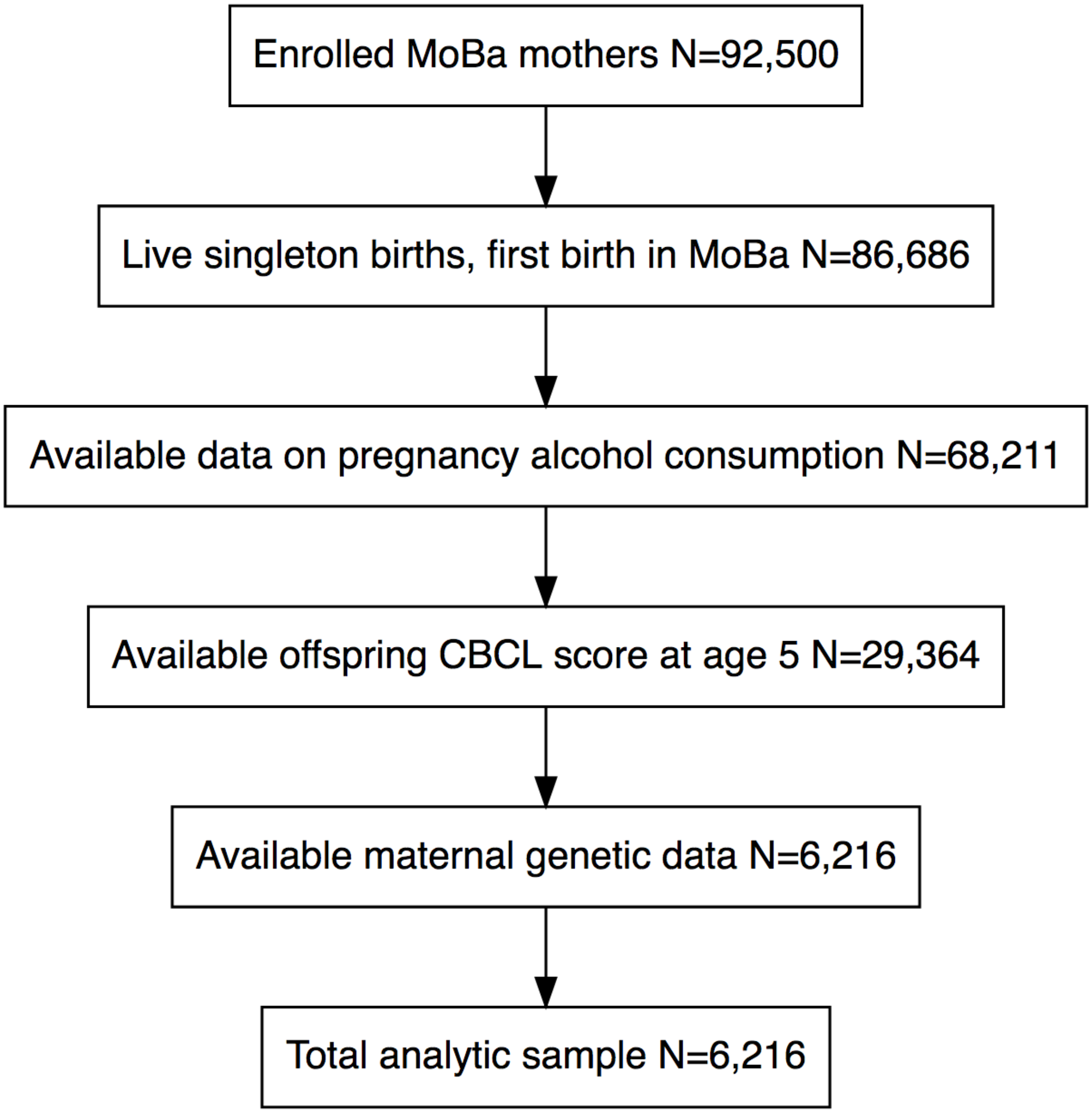

Supplementary figure 3: Cropped visualization of the application of the instrumental inequalities to models for the average causal effect of any vs. no alcohol consumption during pregnancy of offspring ADHD in the Avon Longitudinal Study of Parents and Children.

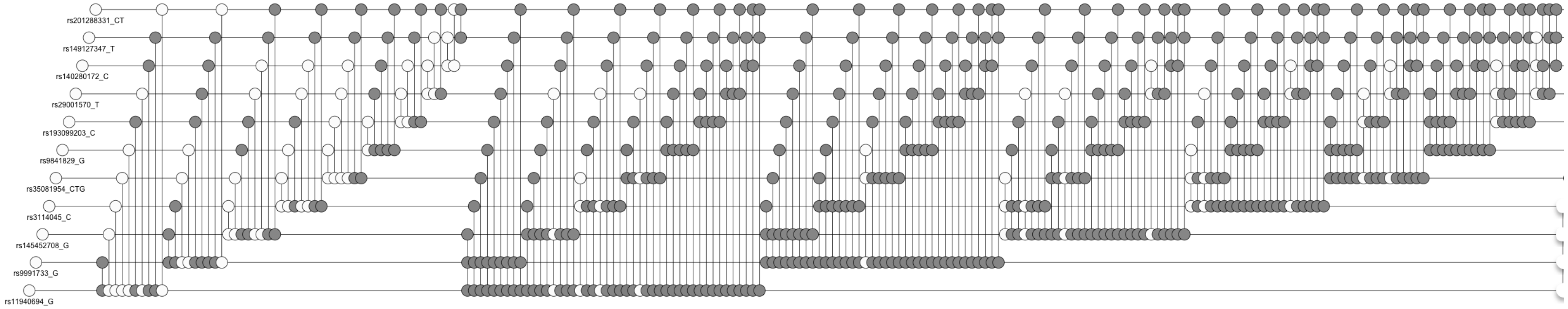


This visualization is cropped such that sets of proposed instruments not shown violated the instrumental inequalities.

Supplementary figure 4: Cropped visualization of the application of the instrumental inequalities to models for the average causal effect of any vs. no alcohol consumption during pregnancy on offspring ADHD in the Norwegian Mother, Father, and Child Study.

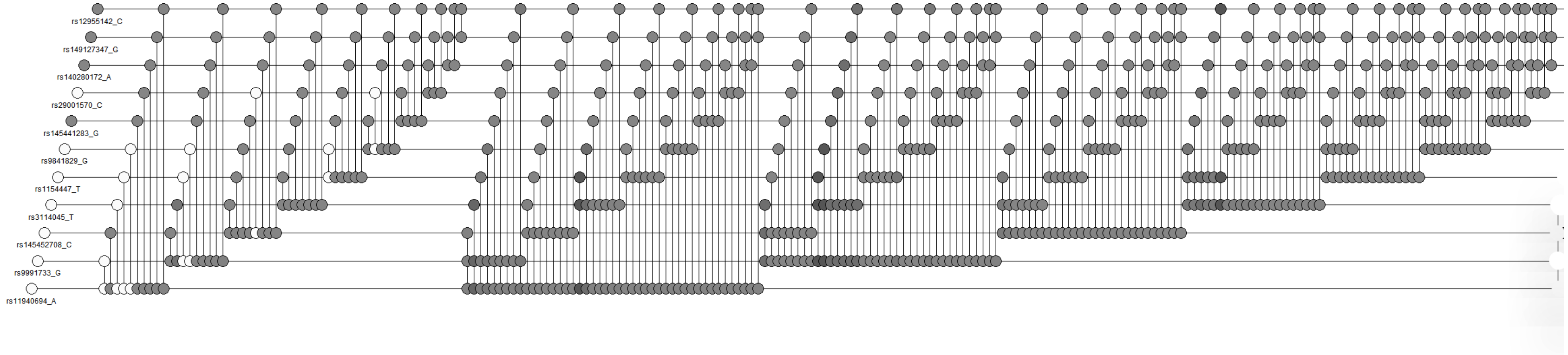


This visualization has been cropped such that all sets of proposed instruments not shown violated the instrumental inequalities.

Supplementary figure 5: Cropped visualization of the application of the instrumental inequalities to models for the average causal effect of moderate vs. no alcohol consumption during pregnancy on offspring ADHD in the Avon Longitudinal Study of Parents and Children.

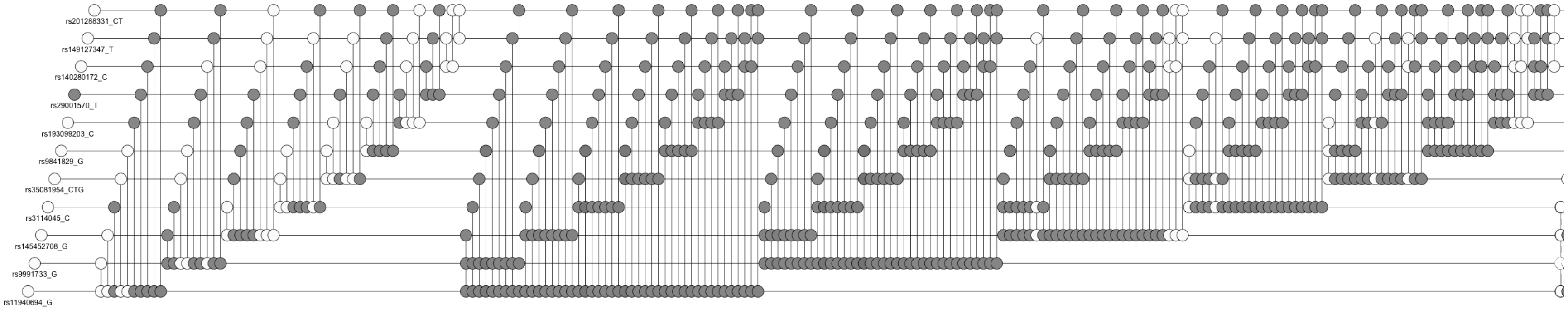


This visualization has been cropped such that all sets of proposed instruments not shown violated the instrumental inequalities.



Supplementary figure 6: Cropped visualization of the application of the instrumental inequalities to models for the average causal effect of moderate vs. no alcohol consumption during pregnancy on offspring ADHD in the Norwegian Mother, Father, and Child Study.

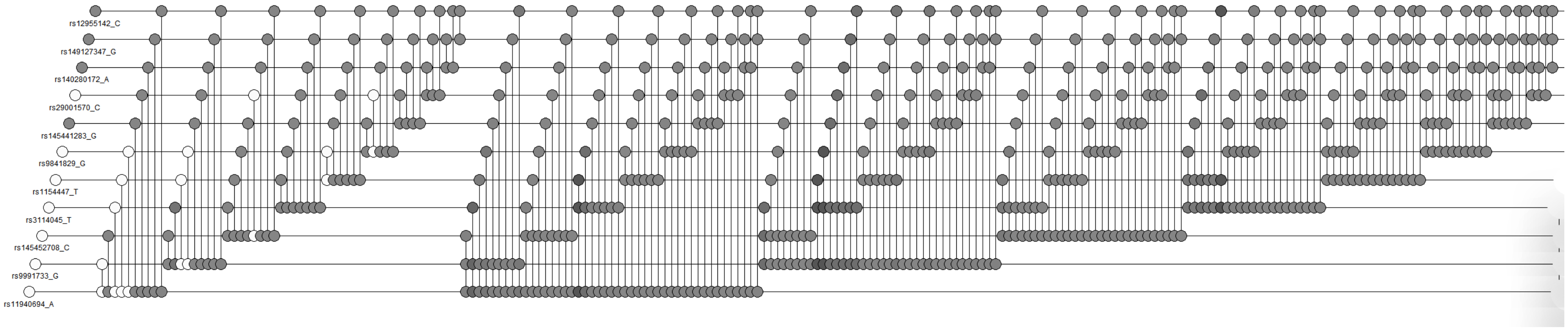


This visualization has been cropped such that all sets of proposed instruments not shown violated the instrumental inequalities.

Supplementary figure 7: Cropped visualization of the application of the instrumental inequalities to models for the average causal effect of alcohol consumption during pregnancy on offspring ADHD in the Avon Longitudinal Study of Parents and Children, grouping alcohol consumption into 3 categories.

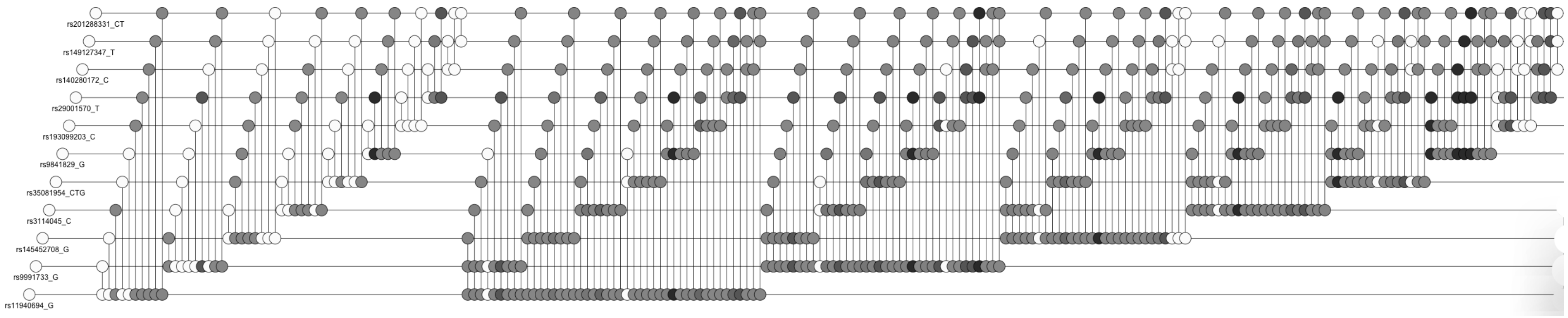


This visualization has been cropped such that all sets of proposed instruments not shown violated the instrumental inequalities.

Supplementary figure 8: Cropped visualization of the application of the instrumental inequalities to models for the average causal effect of alcohol consumption during pregnancy on offspring ADHD in the Avon Longitudinal Study of Parents and Children, grouping alcohol consumption into 4 categories.

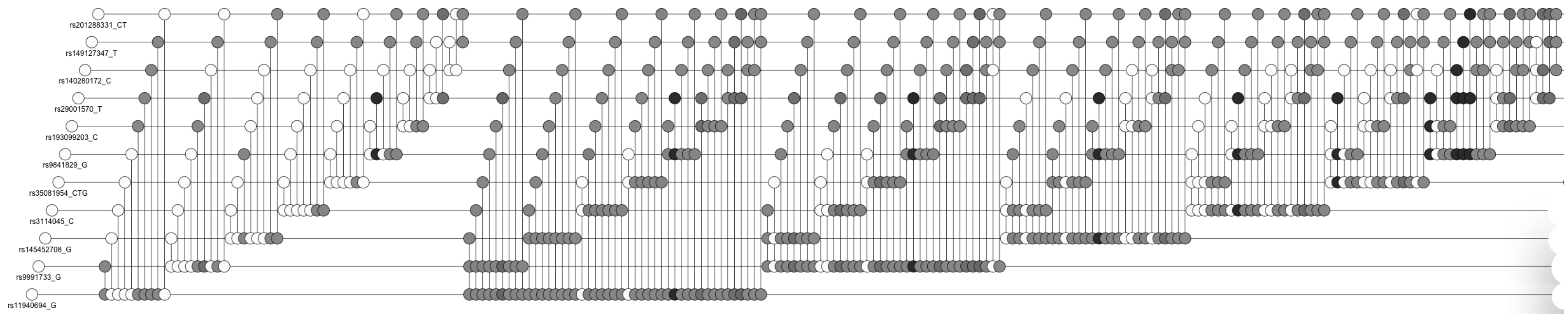


This visualization has been cropped such that all sets of proposed instruments not shown violated the instrumental inequalities.

Supplementary figure 9: Cropped visualization of the application of the instrumental inequalities to models for the average causal effect of alcohol consumption during pregnancy on offspring ADHD in the Avon Longitudinal Study of Parents and Children, grouping alcohol consumption into 7 categories.

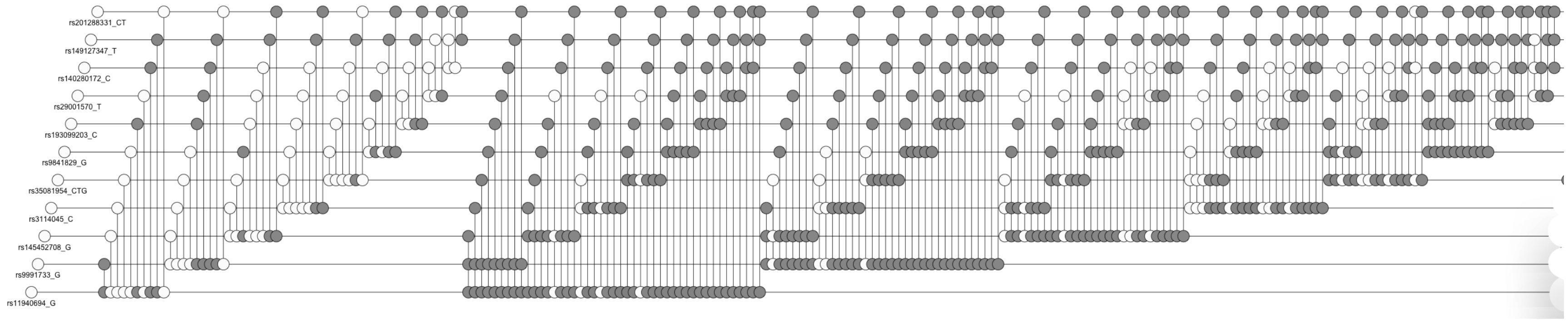


This visualization has been cropped such that all sets of proposed instruments not shown violated the instrumental inequalities.

Supplementary figure 10: Cropped visualization of the application of the instrumental inequalities to models for the average causal effect of alcohol consumption during pregnancy on offspring ADHD in the Norwegian Mother, Father, and Child Study, grouping alcohol consumption into 3 categories.

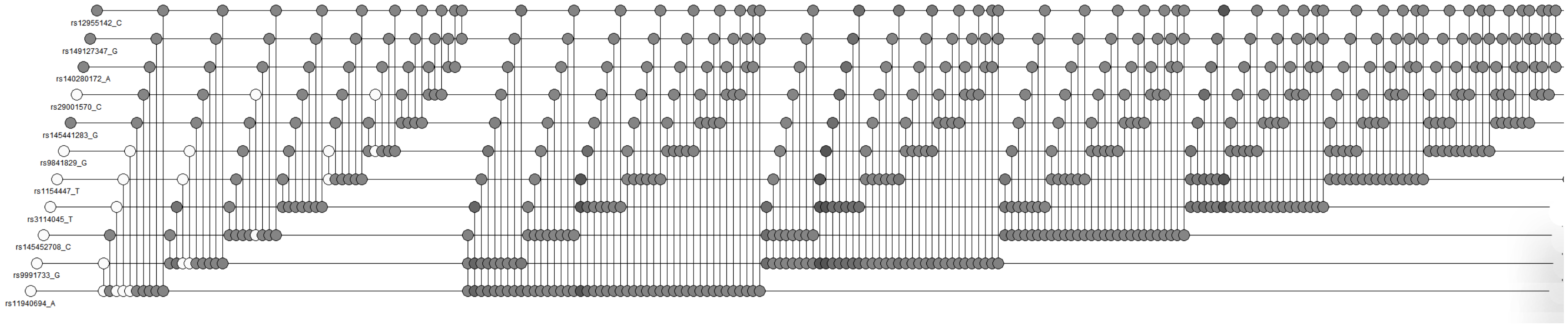


This visualization has been cropped such that all sets of proposed instruments not shown violated the instrumental inequalities.

Supplementary figure 11: Cropped visualization of the application of the instrumental inequalities to models for the average causal effect of alcohol consumption during pregnancy on offspring ADHD in the Norwegian Mother, Father, and Child Study, grouping alcohol consumption into 4 categories.

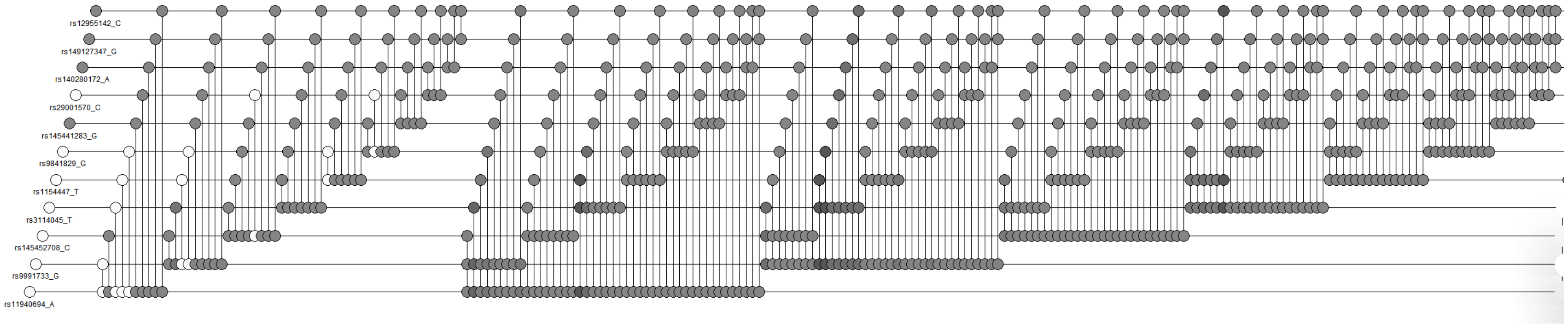


This visualization has been cropped such that all sets of proposed instruments not shown violated the instrumental inequalities.

Supplementary figure 12: Cropped visualization of the application of the instrumental inequalities to models for the average causal effect of alcohol consumption during pregnancy on offspring ADHD in the Norwegian Mother, Father, and Child Study, grouping alcohol consumption into 7 categories.

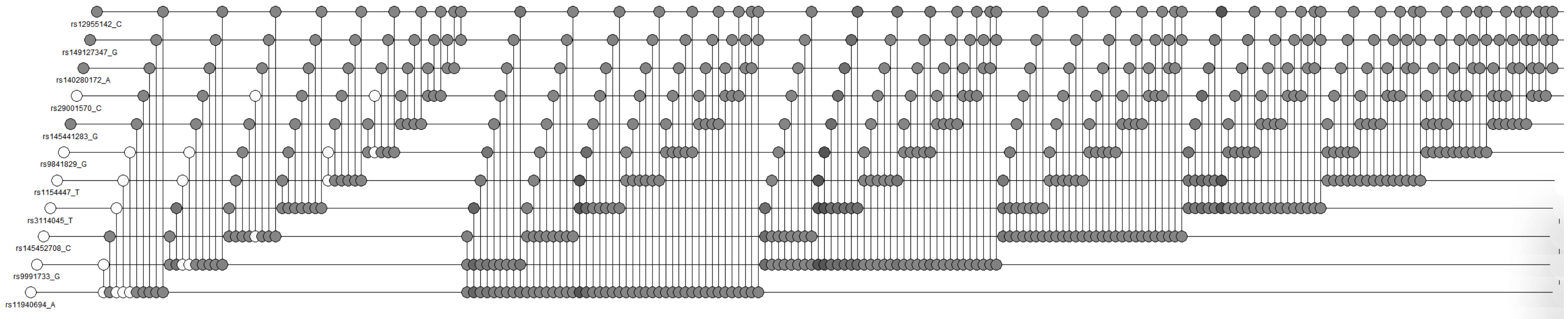


This visualization has been cropped such that all sets of proposed instruments not shown violated the instrumental inequalities.

Supplementary figure 13: Cropped visualization of the application of the instrumental inequalities to models for the average causal effect of any vs. no alcohol consumption during pregnancy on offspring ADHD in the Avon Longitudinal Study of Parents and Children, inverse probability weighted for 10 principal components.

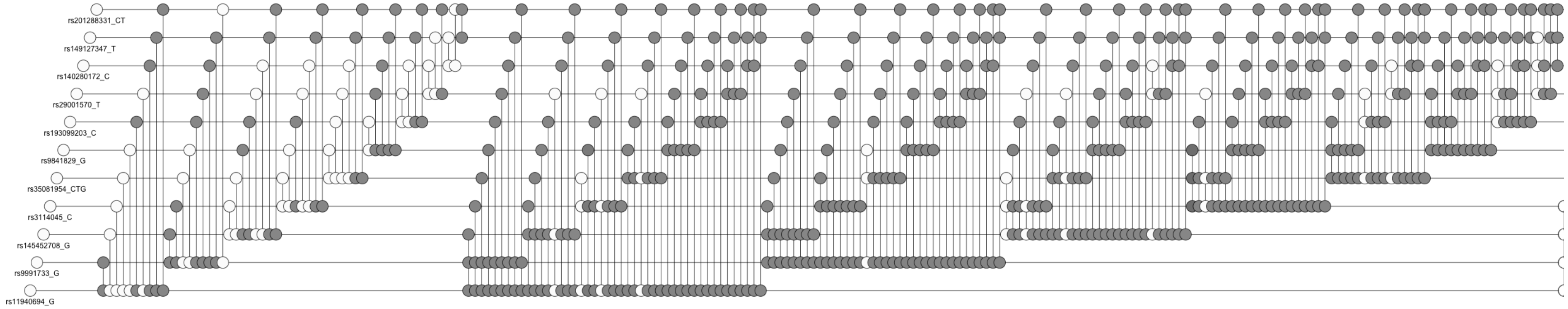


This visualization has been cropped such that all sets of proposed instruments not shown violated the instrumental inequalities.

Supplementary figure 14: Cropped visualization of the application of the instrumental inequalities to models for the average causal effect of moderate vs. no alcohol consumption during pregnancy on offspring ADHD in the Avon Longitudinal Study of Parents and Children, inverse probability weighted for 10 principal components.

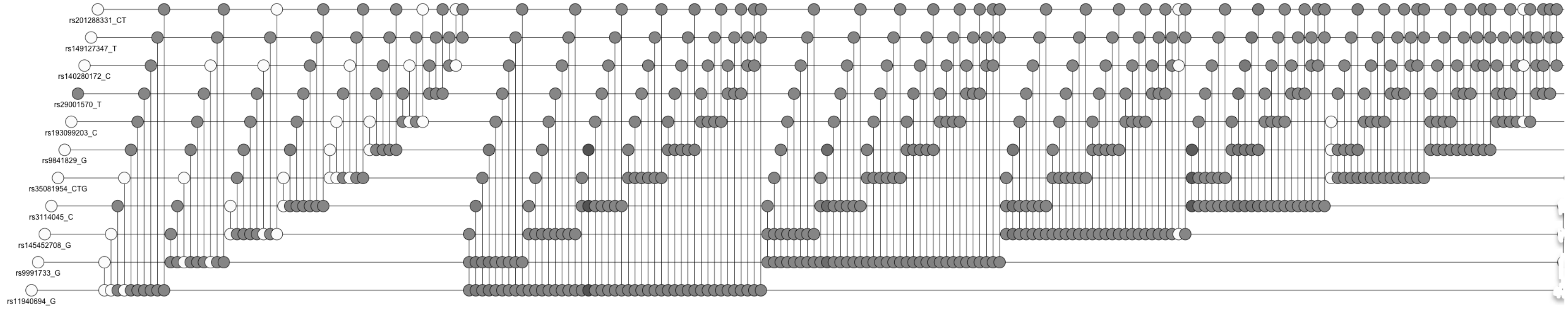


This visualization has been cropped such that all sets of proposed instruments not shown violated the instrumental inequalities.

Supplementary figure 15: Cropped visualization of the application of the instrumental inequalities to models for the average causal effect of any vs. no alcohol consumption during pregnancy on offspring ADHD in the Norwegian Mother, Father, and Child Study, inverse probability weighted for 10 principal components.

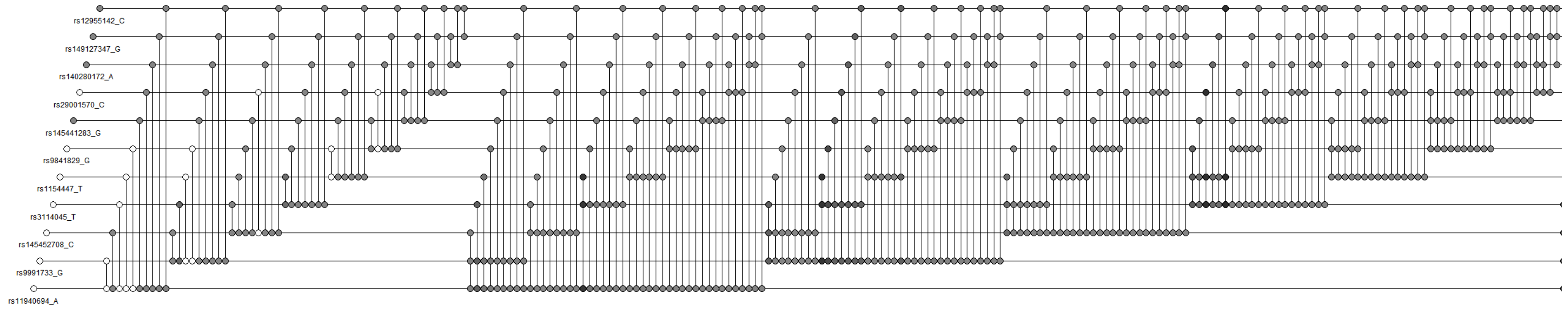


This visualization has been cropped such that all sets of proposed instruments not shown violated the instrumental inequalities.

Supplementary figure 16: Cropped visualization of the application of the instrumental inequalities to models for the average causal effect of moderate vs. no alcohol consumption during pregnancy on offspring ADHD in the Norwegian Mother, Father, and Child Study, inverse probability weighted for 10 principal components.

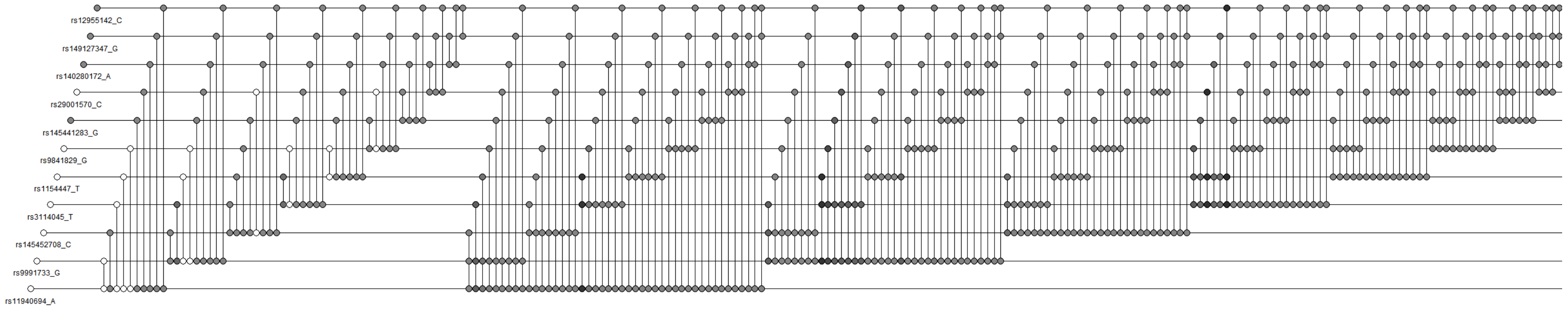


This visualization has been cropped such that all sets of proposed instruments not shown violated the instrumental inequalities.

Supplementary figure 17: Bounds on the average causal effect of any vs. no alcohol consumption on offspring ADHD in the Avon Longitudinal Study of Parents and Children, IP weighted for 10 principal components.

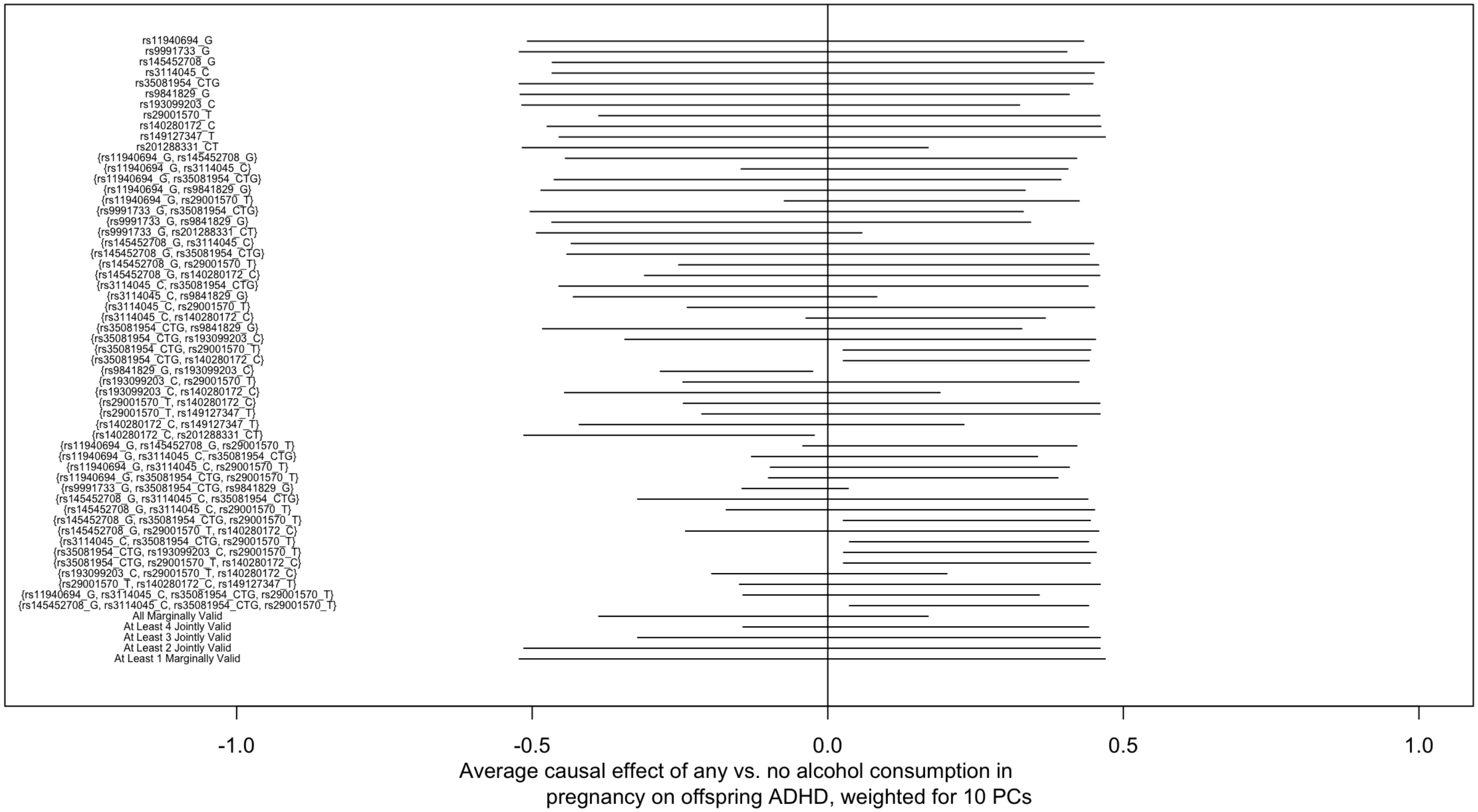

Supplementary figure 18: Bounds on the average causal effect of any vs. no alcohol consumption on offspring ADHD in the Norwegian Mother, Father, and Child Study, IP weighted for 10 principal components.

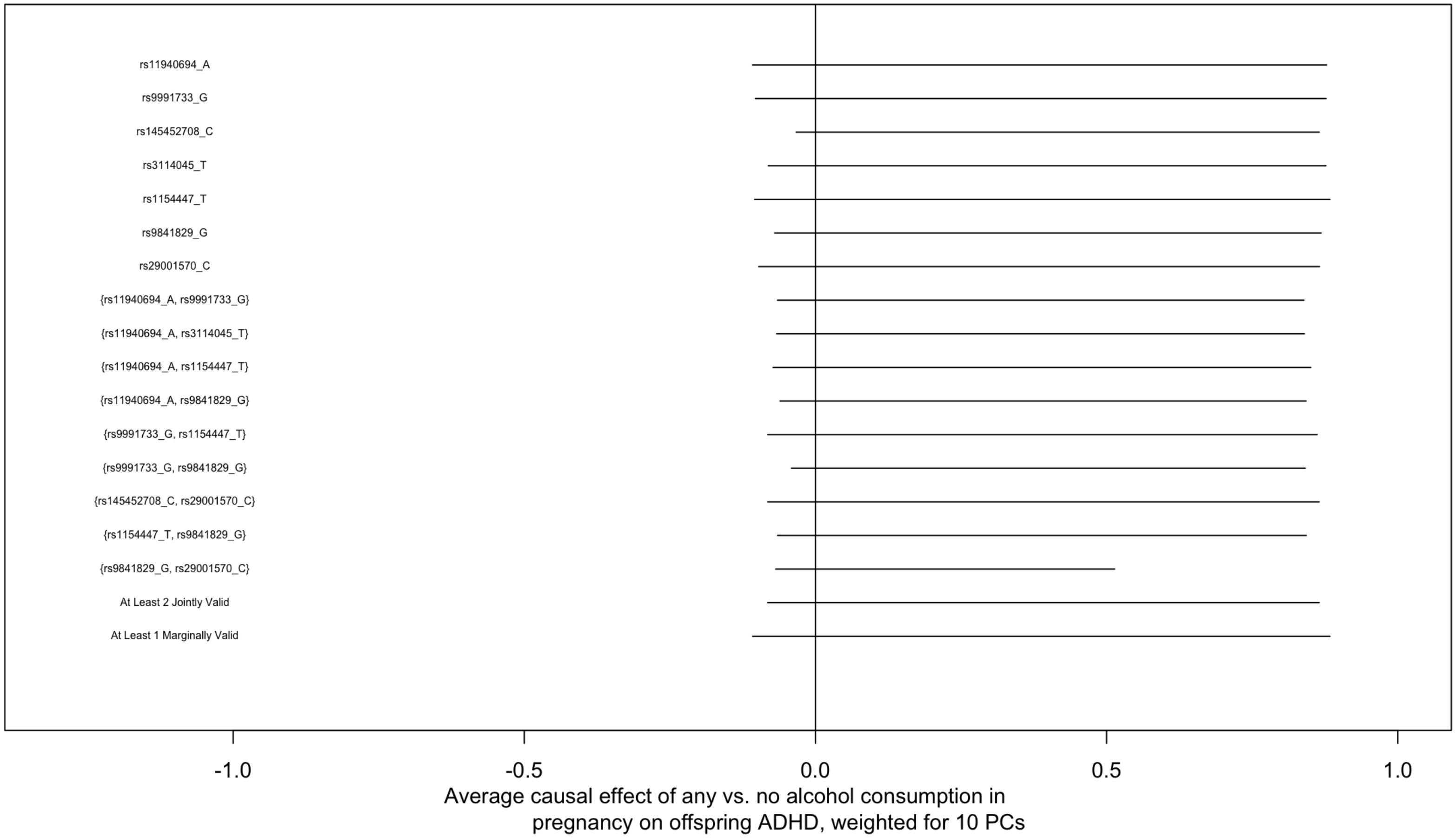

Supplementary figure 19: Bounds on the average causal effect of moderate vs. no alcohol consumption on offspring ADHD in the Avon Longitudinal Study of Parents and Children, IP weighted for 10 principal components.

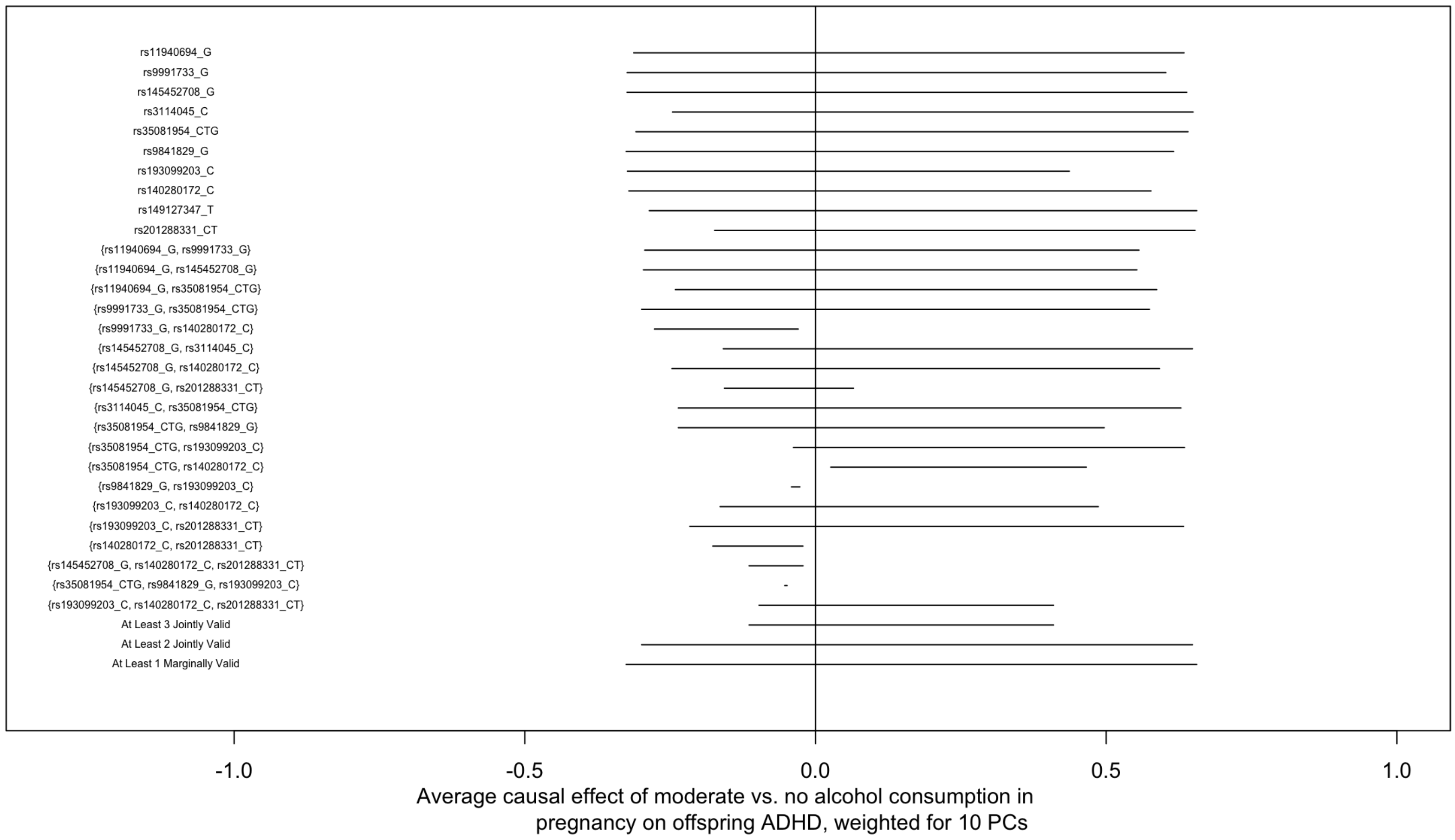

Supplementary figure 20: Bounds on the average causal effect of moderate vs. no alcohol consumption on offspring ADHD in the Norwegian Mother, Father, and Child Study, IP weighted for 10 principal components.

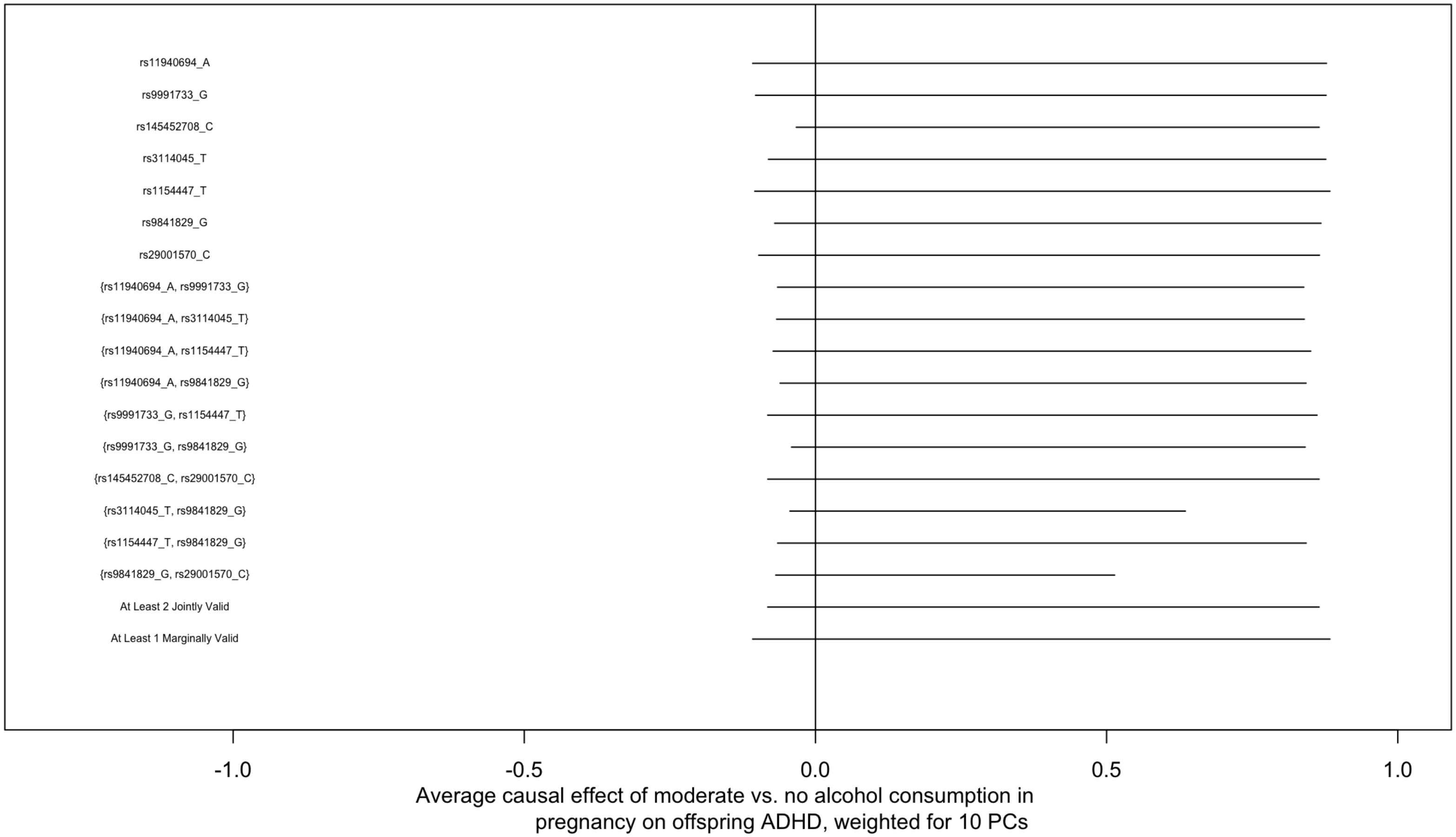

### 9 Supplementary tables 1-9

Supplementary table 1: Bounds and point estimates for the average causal effect of any vs. no alcohol consumption during pregnancy on offspring ADHD in the Avon Longitudinal Study of Parents and Children

| Proposed Instruments | Lower Bound | Upper Bound | Point Estimate (95% CI) |
|---|---|---|---|
| rs11940694_G | -0.51 | 0.43 | 0.06 (-0.19, 0.31) |
| rs9991733_G | -0.52 | 0.41 | 0.02 (-0.59, 0.53) |
| rs145452708_G | -0.47 | 0.47 | 0.11 (-2.22, 2.06) |
| rs3114045_C | -0.48 | 0.45 | 0.02 (-0.37, 0.39) |
| rs35081954_CTG | -0.52 | 0.45 | -0.31 (-1.46, 0.51) |
| rs9841829_G | -0.52 | 0.42 | -0.03 (-0.75, 0.61) |
| rs193099203_C | -0.52 | 0.33 | -0.16 (-2.17, 1.19) |
| rs29001570_T | -0.39 | 0.46 | 0.14 (-1.11, 1.18) |
| rs140280172_C | -0.47 | 0.46 | 0.14 (-1.11, 1.18) |
| rs149127347_T | -0.45 | 0.47 | -1.7 (-9.31, 0.34) |
| rs201288331_CT | -0.51 | 0.32 | 0 (-0.6, 0.58) |
| c("rs11940694_G", "rs145452708_G") | -0.41 | 0.42 | 0.09 (-0.05, 0.25) |
| c("rs11940694_G", "rs3114045_C") | -0.15 | 0.40 | 0.07 (-0.05, 0.2) |
| c("rs11940694_G", "rs35081954_CTG") | -0.46 | 0.38 | 0.06 (-0.06, 0.23) |
| c("rs11940694_G", "rs9841829_G") | -0.48 | 0.36 | 0.04 (-0.1, 0.23) |
| c("rs11940694_G", "rs29001570_T") | -0.49 | 0.42 | 0.08 (-0.13, 0.29) |
| c("rs11940694_G", "rs201288331_CT") | -0.32 | 0.14 | 0.02 (-0.11, 0.17) |
| c("rs9991733_G", "rs35081954_CTG") | -0.50 | 0.33 | -0.05 (-0.31, 0.11) |
| c("rs9991733_G", "rs9841829_G") | -0.44 | 0.31 | 0.15 (-0.02, 0.44) |
| c("rs9991733_G", "rs201288331_CT") | -0.31 | 0.15 | -0.07 (-0.22, 0.05) |
| c("rs145452708_G", "rs3114045_C") | -0.38 | 0.45 | 0.02 (-0.19, 0.25) |
| c("rs145452708_G", "rs35081954_CTG") | -0.43 | 0.44 | -0.03 (-0.45, 0.31) |
| c("rs145452708_G", "rs29001570_T") | -0.39 | 0.46 | 0.09 (-0.33, 0.57) |
| c("rs145452708_G", "rs140280172_C") | -0.37 | 0.46 | 0.04 (-0.22, 0.45) |
| c("rs3114045_C", "rs35081954_CTG") | -0.47 | 0.44 | 0.12 (-0.19, 0.53) |
| c("rs3114045_C", "rs9841829_G") | -0.38 | 0.39 | 0.02 (-0.14, 0.19) |
| c("rs3114045_C", "rs29001570_T") | -0.39 | 0.45 | 0.09 (-0.14, 0.46) |
| c("rs3114045_C", "rs140280172_C") | -0.32 | 0.45 | 0.03 (-0.13, 0.28) |
| c("rs35081954_CTG", "rs9841829_G") | -0.48 | 0.37 | -0.2 (-0.65, -0.09) |
| c("rs35081954_CTG", "rs193099203_C") | -0.50 | 0.32 | -0.27 (-0.84, -0.12) |

Supplementary table 1 contd:

| Proposed Instruments | Lower Bound | Upper Bound | Point Estimate (95% CI) |
|---|---|---|---|
| c(“rs35081954_CTG”, “rs29001570_T”) | 0.03 | 0.44 | -0.07 (-0.48, 0.28) |
| c(“rs35081954_CTG”, “rs140280172_C”) | 0.03 | 0.44 | -0.05 (-0.35, 0.22) |
| c(“rs9841829_G”, “rs193099203_C”) | -0.50 | -0.03 | -0.06 (-0.29, 0.11) |
| c(“rs193099203_C”, “rs29001570_T”) | -0.39 | 0.35 | -0.06 (-0.37, 0.17) |
| c(“rs193099203_C”, “rs140280172_C”) | -0.49 | 0.21 | -0.06 (-0.24, 0.03) |
| c(“rs29001570_T”, “rs140280172_C”) | -0.39 | 0.46 | 0.26 (-0.01, 0.91) |
| c(“rs29001570_T”, “rs149127347_T”) | -0.35 | 0.46 | -0.13 (-1.02, 0.75) |
| c(“rs140280172_C”, “rs149127347_T”) | -0.37 | 0.46 | 0.36 (-0.16, 1.27) |
| c(“rs140280172_C”, “rs201288331_CT”) | -0.45 | -0.02 | 0.07 (-0.18, 0.34) |
| c(“rs11940694_G”, “rs145452708_G”, “rs29001570_T”) | -0.41 | 0.42 | 0.11 (0, 0.26) |
| c(“rs11940694_G”, “rs3114045_C”, “rs35081954_CTG”) | -0.14 | 0.36 | 0.07 (-0.01, 0.21) |
| c(“rs11940694_G”, “rs3114045_C”, “rs29001570_T”) | -0.18 | 0.41 | 0.09 (-0.01, 0.26) |
| c(“rs11940694_G”, “rs35081954_CTG”, “rs29001570_T”) | -0.46 | 0.38 | 0.09 (-0.04, 0.26) |
| c(“rs9991733_G”, “rs35081954_CTG”, “rs9841829_G”) | -0.30 | 0.17 | -0.01 (-0.12, 0.09) |
| c(“rs145452708_G”, “rs3114045_C”, “rs35081954_CTG”) | -0.37 | 0.44 | 0.16 (0.02, 0.5) |
| c(“rs145452708_G”, “rs3114045_C”, “rs29001570_T”) | -0.23 | 0.45 | 0.05 (-0.06, 0.23) |
| c(“rs145452708_G”, “rs35081954_CTG”, “rs29001570_T”) | 0.03 | 0.44 | 0.02 (-0.2, 0.28) |
| c(“rs145452708_G”, “rs29001570_T”, “rs140280172_C”) | -0.37 | 0.46 | 0.04 (-0.13, 0.31) |
| c(“rs3114045_C”, “rs35081954_CTG”, “rs9841829_G”) | -0.35 | 0.32 | -0.01 (-0.17, 0.15) |
| c(“rs3114045_C”, “rs35081954_CTG”, “rs29001570_T”) | 0.04 | 0.44 | 0.18 (0, 0.61) |
| c(“rs3114045_C”, “rs29001570_T”, “rs140280172_C”) | 0.02 | 0.45 | 0.07 (-0.01, 0.28) |
| c(“rs35081954_CTG”, “rs193099203_C”, “rs29001570_T”) | 0.03 | 0.33 | -0.1 (-0.41, 0.08) |
| c(“rs35081954_CTG”, “rs29001570_T”, “rs140280172_C”) | 0.03 | 0.44 | 0 (-0.19, 0.21) |
| c(“rs193099203_C”, “rs29001570_T”, “rs140280172_C”) | -0.39 | 0.22 | -0.01 (-0.11, 0.07) |
| c(“rs29001570_T”, “rs140280172_C”, “rs149127347_T”) | -0.29 | 0.46 | 0.07 (-0.37, 0.46) |
| c(“rs11940694_G”, “rs3114045_C”, “rs35081954_CTG”, “rs29001570_T”) | -0.17 | 0.37 | 0.09 (0.01, 0.24) |
| c(“rs145452708_G”, “rs3114045_C”, “rs35081954_CTG”, “rs29001570_T”) | 0.04 | 0.44 | 0.16 (0.03, 0.43) |
| At Least 4 Jointly Valid | -0.17 | 0.44 | NA |
| At Least 3 Jointly Valid | -0.46 | 0.46 | NA |
| At Least 2 Jointly Valid | -0.50 | 0.46 | NA |
| At Least 1 Marginally Valid | -0.52 | 0.47 | NA |

Supplementary table 2: Bounds and point estimates for the average causal effect of any vs. no alcohol consumption during pregnancy on offspring ADHD in the Norwegian Mother and Child Study

| Proposed Instruments | Lower Bound | Upper Bound | Point Estimate (95% CI) |
|---|---|---|---|
| rs11940694_G | -0.11 | 0.88 | -0.71 (-2.74, 0.81) |
| rs9991733_G | -0.10 | 0.87 | -0.32 (-2.68, 1.43) |
| rs145452708_G | -0.03 | 0.87 | 0.49 (-1.9, 3.28) |
| rs3114045_C | -0.09 | 0.88 | 0.24 (-2.95, 3.38) |
| rs35081954_CTG | -0.10 | 0.88 | 0.13 (-1.06, 1.5) |
| rs9841829_G | -0.08 | 0.87 | 0.3 (-0.37, 0.79) |
| rs29001570_T | -0.11 | 0.86 | -0.94 (-10.64, 5.47) |
| c("rs11940694_G", "rs9991733_G") | -0.08 | 0.83 | -0.47 (-1.1, -0.2) |
| c("rs11940694_G", "rs3114045_C") | -0.09 | 0.83 | -0.57 (-1.63, -0.43) |
| c("rs11940694_G", "rs35081954_CTG") | -0.08 | 0.85 | 0.05 (-0.35, 0.47) |
| c("rs11940694_G", "rs9841829_G") | -0.06 | 0.84 | -0.04 (-0.33, 0.27) |
| c("rs9991733_G", "rs35081954_CTG") | -0.09 | 0.86 | 0.04 (-0.38, 0.54) |
| c("rs9991733_G", "rs9841829_G") | -0.04 | 0.83 | 0.21 (-0.05, 0.58) |
| c("rs145452708_G", "rs29001570_T") | -0.03 | 0.86 | 0.02 (-1.26, 0.97) |
| c("rs35081954_CTG", "rs9841829_G") | -0.06 | 0.84 | 0.22 (-0.03, 0.66) |
| c("rs9841829_G", "rs29001570_T") | -0.07 | 0.73 | 0.17 (-0.15, 0.47) |
| At Least 2 Jointly Valid | -0.09 | 0.86 | NA |
| At Least 1 Marginally Valid | -0.11 | 0.88 | NA |

Supplementary table 3: Bounds and point estimates for the average causal effect of moderate vs. no alcohol consumption during pregnancy on offspring ADHD in the Avon Longitudinal Study of Parents and Children

| Proposed Instruments | Lower Bound | Upper Bound | Point Estimate (95% CI) |
|---|---|---|---|
| rs11940694_G | -0.31 | 0.63 | 0.06 (-0.33, 0.39) |
| rs9991733_G | -0.32 | 0.61 | 0.12 (-0.95, 1.55) |
| rs145452708_G | -0.32 | 0.64 | -0.09 (-3.96, 2.39) |
| rs3114045_C | -0.26 | 0.65 | 0.21 (-0.29, 0.88) |
| rs35081954_CTG | -0.31 | 0.64 | -0.17 (-0.64, 0.24) |
| rs9841829_G | -0.33 | 0.64 | 0.03 (-0.75, 1.15) |
| rs193099203_C | -0.32 | 0.44 | -0.09 (-0.97, 0.7) |
| rs140280172_C | -0.32 | 0.58 | -0.24 (-3.18, 1.99) |
| rs149127347_T | -0.29 | 0.66 | 0.61 (-3.08, 6.3) |
| rs201288331_CT | -0.24 | 0.65 | 0.12 (-0.36, 0.75) |
| c("rs11940694_G", "rs9991733_G") | -0.28 | 0.56 | -0.01 (-0.25, 0.18) |
| c("rs11940694_G", "rs145452708_G") | -0.30 | 0.52 | 0 (-0.21, 0.17) |
| c("rs11940694_G", "rs35081954_CTG") | -0.24 | 0.60 | 0 (-0.16, 0.15) |
| c("rs11940694_G", "rs9841829_G") | -0.25 | 0.57 | 0.06 (-0.11, 0.29) |
| c("rs9991733_G", "rs35081954_CTG") | -0.29 | 0.55 | -0.09 (-0.36, 0.09) |
| c("rs9991733_G", "rs9841829_G") | -0.11 | 0.47 | 0.2 (0.05, 0.59) |
| c("rs9991733_G", "rs140280172_C") | -0.21 | -0.03 | -0.01 (-0.17, 0.14) |
| c("rs145452708_G", "rs3114045_C") | -0.23 | 0.62 | 0.13 (-0.07, 0.43) |
| c("rs145452708_G", "rs140280172_C") | -0.22 | 0.56 | -0.18 (-0.67, -0.02) |
| c("rs145452708_G", "rs149127347_T") | -0.19 | 0.63 | 0.3 (-0.42, 1.02) |
| c("rs145452708_G", "rs201288331_CT") | -0.23 | 0.32 | 0 (-0.26, 0.21) |
| c("rs3114045_C", "rs35081954_CTG") | -0.25 | 0.63 | -0.13 (-0.56, 0.14) |
| c("rs3114045_C", "rs9841829_G") | -0.20 | 0.57 | 0.02 (-0.15, 0.25) |
| c("rs3114045_C", "rs149127347_T") | 0.10 | 0.59 | 0.25 (-0.08, 0.59) |
| c("rs35081954_CTG", "rs9841829_G") | -0.28 | 0.59 | -0.2 (-0.53, -0.09) |
| c("rs35081954_CTG", "rs193099203_C") | -0.29 | 0.42 | -0.13 (-0.36, 0.04) |
| c("rs35081954_CTG", "rs140280172_C") | 0.03 | 0.55 | -0.16 (-0.42, 0) |
| c("rs35081954_CTG", "rs149127347_T") | 0.17 | 0.53 | -0.11 (-0.7, 0.08) |
| c("rs9841829_G", "rs193099203_C") | -0.32 | -0.03 | -0.07 (-0.29, 0.13) |
| c("rs193099203_C", "rs140280172_C") | -0.28 | 0.32 | -0.06 (-0.19, 0.01) |
| c("rs193099203_C", "rs149127347_T") | -0.32 | 0.32 | -0.09 (-0.26, -0.02) |
| c("rs193099203_C", "rs201288331_CT") | -0.24 | 0.56 | 0.01 (-0.24, 0.29) |

Supplementary table 3 contd:

| Proposed Instruments | Lower Bound | Upper Bound | Point Estimate (95% CI) |
|---|---|---|---|
| c("rs140280172_C", "rs149127347_T") | -0.19 | 0.54 | 0.08 (-0.55, 0.41) |
| c("rs140280172_C", "rs201288331_CT") | -0.24 | -0.02 | -0.03 (-0.29, 0.13) |
| c("rs149127347_T", "rs201288331_CT") | 0.08 | 0.65 | 0.09 (-0.59, 0.5) |
| c("rs145452708_G", "rs3114045_C", "rs149127347_T") | 0.11 | -0.11 | 0.1 (-0.22, 0.3) |
| c("rs145452708_G", "rs140280172_C", "rs149127347_T") | 0.25 | 0.53 | 0.18 (-0.37, 0.47) |
| c("rs145452708_G", "rs140280172_C", "rs201288331_CT") | -0.09 | -0.04 | -0.08 (-0.27, 0.06) |
| c("rs145452708_G", "rs149127347_T", "rs201288331_CT") | 0.20 | -0.20 | 0.08 (-0.33, 0.26) |
| c("rs3114045_C", "rs35081954_CTG", "rs9841829_G") | -0.13 | 0.49 | -0.18 (-0.43, -0.1) |
| c("rs3114045_C", "rs35081954_CTG", "rs149127347_T") | 0.25 | 0.32 | -0.1 (-0.5, 0.01) |
| c("rs35081954_CTG", "rs9841829_G", "rs193099203_C") | -0.28 | -0.04 | -0.13 (-0.33, -0.05) |
| c("rs35081954_CTG", "rs193099203_C", "rs149127347_T") | -0.29 | -0.03 | -0.11 (-0.27, -0.01) |
| c("rs35081954_CTG", "rs140280172_C", "rs149127347_T") | 0.33 | 0.32 | 0.05 (-0.32, 0.22) |
| c("rs193099203_C", "rs140280172_C", "rs149127347_T") | 0.02 | -0.02 | -0.03 (-0.11, -0.01) |
| c("rs193099203_C", "rs140280172_C", "rs201288331_CT") | -0.24 | 0.49 | 0.01 (-0.13, 0.18) |
| c("rs193099203_C", "rs149127347_T", "rs201288331_CT") | 0.02 | 0.49 | 0.01 (-0.15, 0.18) |
| c("rs140280172_C", "rs149127347_T", "rs201288331_CT") | 0.12 | -0.12 | 0.02 (-0.41, 0.2) |
| c("rs145452708_G", "rs140280172_C", "rs149127347_T", "rs201288331_CT") | 0.50 | -0.50 | 0.09 (-0.3, 0.22) |
| c("rs193099203_C", "rs140280172_C", "rs149127347_T", "rs201288331_CT") | 0.02 | 0.32 | 0.01 (-0.13, 0.12) |
| At Least 4 Jointly Valid | 0.02 | 0.32 | NA |
| At Least 3 Jointly Valid | -0.29 | 0.53 | NA |
| At Least 2 Jointly Valid | -0.32 | 0.65 | NA |
| At Least 1 Marginally Valid | -0.33 | 0.66 | NA |

Supplementary table 4: Bounds and point estimates for the average causal effect of moderate vs. no alcohol consumption during pregnancy on offspring ADHD in the Norwegian Mother, Father, and Child Study

| Proposed Instruments | Lower Bound | Upper Bound | Point Estimate (95% CI) |
|---|---|---|---|
| rs11940694_G | -0.11 | 0.88 | -0.71 (-2.85, 0.63) |
| rs9991733_G | -0.10 | 0.87 | -0.32 (-2.63, 1.66) |
| rs145452708_G | -0.03 | 0.87 | 0.49 (-2.14, 3.48) |
| rs3114045_C | -0.09 | 0.88 | 0.24 (-2.56, 3.33) |
| rs35081954_CTG | -0.10 | 0.88 | 0.13 (-1.19, 1.7) |
| rs9841829_G | -0.08 | 0.87 | 0.3 (-0.36, 0.78) |
| rs29001570_T | -0.11 | 0.86 | -0.94 (-8.66, 5.33) |
| c("rs11940694_G", "rs9991733_G") | -0.08 | 0.83 | -0.47 (-1.14, -0.17) |
| c("rs11940694_G", "rs3114045_C") | -0.09 | 0.83 | -0.57 (-1.63, -0.46) |
| c("rs11940694_G", "rs35081954_CTG") | -0.08 | 0.85 | 0.05 (-0.3, 0.43) |
| c("rs11940694_G", "rs9841829_G") | -0.06 | 0.84 | -0.04 (-0.31, 0.25) |
| c("rs9991733_G", "rs35081954_CTG") | -0.09 | 0.86 | 0.04 (-0.38, 0.47) |
| c("rs9991733_G", "rs9841829_G") | -0.04 | 0.83 | 0.21 (-0.03, 0.58) |
| c("rs145452708_G", "rs29001570_T") | -0.03 | 0.86 | 0.02 (-0.99, 0.87) |
| c("rs35081954_CTG", "rs9841829_G") | -0.06 | 0.84 | 0.22 (-0.05, 0.67) |
| c("rs9841829_G", "rs29001570_T") | -0.07 | 0.73 | 0.17 (-0.12, 0.49) |
| At Least 2 Jointly Valid | -0.09 | 0.86 | NA |
| At Least 1 Marginally Valid | -0.11 | 0.88 | NA |

Supplementary table 5: Bounds and point estimates for the average causal effect of any vs. no alcohol consumption during pregnancy on offspring ADHD in the Avon Longitudinal Study of Parents and Children, inverse probability weighted for 10 principal components

| Proposed Instruments | Lower Bound | Upper Bound | Point Estimate (95% CI) |
|---|---|---|---|
| rs11940694_G | -0.51 | 0.43 | 0.05 (-0.21, 0.3) |
| rs9991733_G | -0.52 | 0.40 | 0.06 (-0.51, 0.56) |
| rs145452708_G | -0.47 | 0.47 | 0.11 (-1.78, 2.2) |
| rs3114045_C | -0.47 | 0.45 | 0.29 (-0.02, 1.1) |
| rs35081954_CTG | -0.52 | 0.45 | -0.33 (-1.88, 0.47) |
| rs9841829_G | -0.52 | 0.41 | 0.08 (-0.58, 0.95) |
| rs193099203_C | -0.52 | 0.32 | -0.16 (-1.55, 1.15) |
| rs29001570_T | -0.39 | 0.46 | 0.14 (-1.55, 1.15) |
| rs140280172_C | -0.47 | 0.46 | 0.42 (-3.81, 5.03) |
| rs149127347_T | -0.45 | 0.47 | -1.62 (-8.6, 2) |
| rs201288331_CT | -0.52 | 0.17 | -0.06 (-0.33, 0.11) |
| c(“rs11940694_G”, “rs145452708_G”) | -0.44 | 0.42 | 0.2 (0.21, 0.47) |
| c(“rs11940694_G”, “rs3114045_C”) | -0.15 | 0.41 | 0.07 (0.03, 0.13) |
| c(“rs11940694_G”, “rs35081954_CTG”) | -0.46 | 0.39 | 0 (-0.21, 0.13) |
| c(“rs11940694_G”, “rs9841829_G”) | -0.49 | 0.33 | -0.03 (-0.35, 0.2) |
| c(“rs11940694_G”, “rs29001570_T”) | -0.07 | 0.43 | 0.04 (-0.03, 0.16) |
| c(“rs9991733_G”, “rs35081954_CTG”) | -0.50 | 0.33 | -0.03 (-0.22, 0.11) |
| c(“rs9991733_G”, “rs9841829_G”) | -0.47 | 0.34 | 0.29 (0.19, 0.87) |
| c(“rs9991733_G”, “rs201288331_CT”) | -0.49 | 0.06 | -0.06 (-0.1, -0.02) |
| c(“rs145452708_G”, “rs3114045_C”) | -0.43 | 0.45 | 0.21 (0.21, 0.55) |
| c(“rs145452708_G”, “rs35081954_CTG”) | -0.44 | 0.44 | 0.2 (0.1, 0.67) |
| c(“rs145452708_G”, “rs29001570_T”) | -0.25 | 0.46 | 0.07 (-0.11, 0.23) |
| c(“rs145452708_G”, “rs140280172_C”) | -0.31 | 0.46 | 0.05 (-0.06, 0.26) |
| c(“rs3114045_C”, “rs35081954_CTG”) | -0.45 | 0.44 | 0.27 (0.12, 0.84) |
| c(“rs3114045_C”, “rs9841829_G”) | -0.43 | 0.08 | -0.03 (-0.21, 0.03) |
| c(“rs3114045_C”, “rs29001570_T”) | -0.24 | 0.45 | 0.07 (-0.09, 0.21) |
| c(“rs3114045_C”, “rs140280172_C”) | -0.04 | 0.37 | 0.03 (-0.05, 0.1) |
| c(“rs35081954_CTG”, “rs9841829_G”) | -0.48 | 0.33 | -0.11 (-0.45, 0.06) |
| c(“rs35081954_CTG”, “rs193099203_C”) | -0.34 | 0.45 | 0.12 (0.1, 0.34) |
| c(“rs35081954_CTG”, “rs29001570_T”) | 0.03 | 0.44 | 0.04 (-0.01, 0.18) |
| c(“rs35081954_CTG”, “rs140280172_C”) | 0.03 | 0.44 | 0.07 (-0.03, 0.26) |
| c(“rs9841829_G”, “rs193099203_C”) | -0.28 | -0.03 | 0.02 (-0.08, 0.14) |

Supplementary table 5 contd:

| Proposed Instruments | Lower Bound | Upper Bound | Point Estimate (95% CI) |
|---|---|---|---|
| c(“rs193099203_C”, “rs29001570_T”) | -0.24 | 0.42 | 0.04 (-0.13, 0.21) |
| c(“rs193099203_C”, “rs140280172_C”) | -0.45 | 0.19 | -0.01 (-0.09, 0.06) |
| c(“rs29001570_T”, “rs140280172_C”) | -0.24 | 0.46 | 0.06 (-0.18, 0.18) |
| c(“rs29001570_T”, “rs149127347_T”) | -0.21 | 0.46 | 0.05 (-0.48, 0.64) |
| c(“rs140280172_C”, “rs149127347_T”) | -0.42 | 0.23 | -0.03 (-0.53, 0.25) |
| c(“rs140280172_C”, “rs201288331_CT”) | -0.51 | -0.02 | -0.04 (-0.15, 0.05) |
| c(“rs11940694_G”, “rs145452708_G”, “rs29001570_T”) | -0.04 | 0.42 | 0.03 (-0.05, 0.09) |
| c(“rs11940694_G”, “rs3114045_C”, “rs35081954_CTG”) | -0.13 | 0.35 | 0.06 (0.03, 0.13) |
| c(“rs11940694_G”, “rs3114045_C”, “rs29001570_T”) | -0.10 | 0.41 | 0.05 (0.02, 0.08) |
| c(“rs11940694_G”, “rs35081954_CTG”, “rs29001570_T”) | -0.10 | 0.39 | 0.04 (-0.02, 0.15) |
| c(“rs9991733_G”, “rs35081954_CTG”, “rs9841829_G”) | -0.15 | 0.03 | -0.03 (-0.1, 0.04) |
| c(“rs145452708_G”, “rs3114045_C”, “rs35081954_CTG”) | -0.32 | 0.44 | 0.12 (0.07, 0.26) |
| c(“rs145452708_G”, “rs3114045_C”, “rs29001570_T”) | -0.17 | 0.45 | 0.05 (0, 0.13) |
| c(“rs145452708_G”, “rs35081954_CTG”, “rs29001570_T”) | 0.03 | 0.44 | 0.04 (-0.03, 0.11) |
| c(“rs145452708_G”, “rs29001570_T”, “rs140280172_C”) | -0.24 | 0.46 | 0.06 (0.02, 0.16) |
| c(“rs3114045_C”, “rs35081954_CTG”, “rs29001570_T”) | 0.04 | 0.44 | 0.04 (-0.07, 0.16) |
| c(“rs35081954_CTG”, “rs193099203_C”, “rs29001570_T”) | 0.03 | 0.45 | 0.04 (0.03, 0.1) |
| c(“rs35081954_CTG”, “rs29001570_T”, “rs140280172_C”) | 0.03 | 0.44 | 0.03 (-0.01, 0.07) |
| c(“rs193099203_C”, “rs29001570_T”, “rs140280172_C”) | -0.20 | 0.20 | 0.01 (-0.02, 0.04) |
| c(“rs29001570_T”, “rs140280172_C”, “rs149127347_T”) | -0.15 | 0.46 | -0.04 (-0.29, 0.11) |
| c(“rs11940694_G”, “rs3114045_C”, “rs35081954_CTG”, “rs29001570_T”) | -0.14 | 0.36 | 0.06 (0.04, 0.11) |
| c(“rs145452708_G”, “rs3114045_C”, “rs35081954_CTG”, “rs29001570_T”) | 0.04 | 0.44 | 0.05 (0.01, 0.1) |
| At Least 4 Jointly Valid | -0.14 | 0.44 | NA |
| At Least 3 Jointly Valid | -0.32 | 0.46 | NA |
| At Least 2 Jointly Valid | -0.51 | 0.46 | NA |
| At Least 1 Marginally Valid | -0.52 | 0.47 | NA |

Supplementary table 6: Bounds and point estimates for the average causal effect of moderate vs. no alcohol consumption during pregnancy on offspring ADHD in the Avon Longitudinal Study of Parents and Children, inverse probability weighted for 10 principal components

| Proposed Instruments | Lower Bound | Upper Bound | Point Estimate (95% CI) |
|---|---|---|---|
| rs11940694_G | -0.31 | 0.63 | 0.02 (-0.37, 0.4) |
| rs9991733_G | -0.32 | 0.60 | 0.24 (-0.83, 1.62) |
| rs145452708_G | -0.32 | 0.64 | -0.08 (-3.8, 2.58) |
| rs3114045_C | -0.25 | 0.65 | 0.26 (-0.23, 1.38) |
| rs35081954_CTG | -0.31 | 0.64 | -0.23 (-0.87, 0.42) |
| rs9841829_G | -0.33 | 0.62 | 0.24 (-0.69, 1.48) |
| rs193099203_C | -0.32 | 0.44 | -0.09 (-0.8, 0.71) |
| rs140280172_C | -0.32 | 0.58 | -0.24 (-3.7, 2.68) |
| rs149127347_T | -0.29 | 0.66 | 0.61 (-3.52, 5.29) |
| rs201288331_CT | -0.17 | 0.65 | 0.14 (-0.15, 0.61) |
| c("rs11940694_G", "rs9991733_G") | -0.29 | 0.56 | 0.09 (-0.1, 0.4) |
| c("rs11940694_G", "rs145452708_G") | -0.30 | 0.55 | -0.11 (-0.38, -0.04) |
| c("rs11940694_G", "rs35081954_CTG") | -0.24 | 0.59 | -0.07 (-0.31, 0.01) |
| c("rs9991733_G", "rs35081954_CTG") | -0.30 | 0.57 | -0.09 (-0.36, 0.02) |
| c("rs9991733_G", "rs140280172_C") | -0.28 | -0.03 | -0.03 (-0.12, 0.05) |
| c("rs145452708_G", "rs3114045_C") | -0.16 | 0.65 | 0.12 (0.03, 0.32) |
| c("rs145452708_G", "rs140280172_C") | -0.25 | 0.59 | -0.1 (-0.39, 0.02) |
| c("rs145452708_G", "rs201288331_CT") | -0.16 | 0.07 | -0.02 (-0.17, 0.01) |
| c("rs3114045_C", "rs35081954_CTG") | -0.24 | 0.63 | 0.17 (0, 0.68) |
| c("rs35081954_CTG", "rs9841829_G") | -0.24 | 0.50 | 0.03 (-0.17, 0.3) |
| c("rs35081954_CTG", "rs193099203_C") | -0.04 | 0.63 | 0.07 (0.04, 0.19) |
| c("rs35081954_CTG", "rs140280172_C") | 0.03 | 0.47 | 0 (-0.1, 0.14) |
| c("rs9841829_G", "rs193099203_C") | -0.04 | -0.03 | 0.04 (-0.06, 0.18) |
| c("rs193099203_C", "rs140280172_C") | -0.16 | 0.49 | -0.01 (-0.11, 0.07) |
| c("rs193099203_C", "rs201288331_CT") | -0.22 | 0.63 | 0.03 (-0.15, 0.27) |
| c("rs140280172_C", "rs201288331_CT") | -0.18 | -0.02 | -0.04 (-0.16, 0.07) |
| c("rs145452708_G", "rs140280172_C", "rs201288331_CT") | -0.11 | -0.02 | -0.02 (-0.07, 0.01) |
| c("rs35081954_CTG", "rs9841829_G", "rs193099203_C") | -0.05 | -0.05 | 0.03 (-0.06, 0.12) |
| c("rs193099203_C", "rs140280172_C", "rs201288331_CT") | -0.10 | 0.41 | 0 (-0.08, 0.04) |
| At Least 3 Jointly Valid | -0.11 | 0.41 | NA |
| At Least 2 Jointly Valid | -0.30 | 0.65 | NA |
| At Least 1 Marginally Valid | -0.33 | 0.66 | NA |

Supplementary table 7: Bounds and point estimates for the average causal effect of any vs. no alcohol consumption during pregnancy on offspring ADHD in the Norwegian Mother and Child Study, inverse probability weighted for 10 principal components

| Proposed Instruments | Lower Bound | Upper Bound | Point Estimate (95% CI) |
|---|---|---|---|
| rs11940694_G | -0.11 | 0.88 | -0.83 (-2.89, 0.44) |
| rs9991733_G | -0.10 | 0.88 | -0.31 (-2.83, 1.75) |
| rs145452708_G | -0.03 | 0.87 | 0.49 (-2.61, 5.07) |
| rs3114045_C | -0.08 | 0.88 | -0.04 (-1.92, 1.99) |
| rs35081954_CTG | -0.10 | 0.88 | 0.2 (-1.46, 2.07) |
| rs9841829_G | -0.07 | 0.87 | 0.41 (-0.42, 0.8) |
| rs29001570_T | -0.10 | 0.87 | -2.28 (-17.79, 6.09) |
| c("rs11940694_G", "rs9991733_G") | -0.07 | 0.84 | -0.39 (-1.07, 0.47) |
| c("rs11940694_G", "rs3114045_C") | -0.07 | 0.84 | -0.23 (-0.86, 0.25) |
| c("rs11940694_G", "rs35081954_CTG") | -0.07 | 0.85 | 0.18 (-0.18, 0.69) |
| c("rs11940694_G", "rs9841829_G") | -0.06 | 0.84 | 0.22 (-0.01, 0.67) |
| c("rs9991733_G", "rs35081954_CTG") | -0.08 | 0.86 | -0.02 (-0.74, 0.52) |
| c("rs9991733_G", "rs9841829_G") | -0.04 | 0.84 | 0.27 (0, 0.72) |
| c("rs145452708_G", "rs29001570_T") | -0.08 | 0.87 | -0.4 (-2.2, 0.25) |
| c("rs35081954_CTG", "rs9841829_G") | -0.06 | 0.84 | 0.33 (0.03, 0.91) |
| c("rs9841829_G", "rs29001570_T") | -0.07 | 0.51 | -0.06 (-0.17, 0.03) |
| At Least 2 Jointly Valid | -0.08 | 0.87 | NA |
| At Least 1 Marginally Valid | -0.11 | 0.88 | NA |

Supplementary table 8: Bounds and point estimates for the average causal effect of moderate vs. no alcohol consumption during pregnancy on offspring ADHD in the Norwegian Mother and Child Study, inverse probability weighted for 10 principal components

| Proposed Instruments | Lower Bound | Upper Bound | Point Estimate (95% CI) |
|---|---|---|---|
| rs11940694_G | -0.11 | 0.88 | -0.83 (-3.12, 0.35) |
| rs9991733_G | -0.10 | 0.88 | -0.31 (-2.65, 1.26) |
| rs145452708_G | -0.03 | 0.87 | 0.49 (-2.03, 2.9) |
| rs3114045_C | -0.08 | 0.88 | -0.04 (-1.89, 2.22) |
| rs35081954_CTG | -0.10 | 0.88 | 0.2 (-1.24, 1.78) |
| rs9841829_G | -0.07 | 0.87 | 0.41 (-0.34, 0.8) |
| rs29001570_T | -0.10 | 0.87 | -2.28 (-15.35, 3.77) |
| c("rs11940694_G", "rs9991733_G") | -0.07 | 0.84 | -0.39 (-1.08, 0.49) |
| c("rs11940694_G", "rs3114045_C") | -0.07 | 0.84 | -0.23 (-0.83, 0.41) |
| c("rs11940694_G", "rs35081954_CTG") | -0.07 | 0.85 | 0.18 (-0.12, 0.7) |
| c("rs11940694_G", "rs9841829_G") | -0.06 | 0.84 | 0.22 (0.01, 0.67) |
| c("rs9991733_G", "rs35081954_CTG") | -0.08 | 0.86 | -0.02 (-0.78, 0.54) |
| c("rs9991733_G", "rs9841829_G") | -0.04 | 0.84 | 0.27 (-0.02, 0.71) |
| c("rs145452708_G", "rs29001570_T") | -0.08 | 0.87 | -0.4 (-2.48, 0.24) |
| c("rs3114045_C", "rs9841829_G") | -0.04 | 0.64 | -0.06 (-0.53, 0.12) |
| c("rs35081954_CTG", "rs9841829_G") | -0.06 | 0.84 | 0.33 (0.04, 0.96) |
| c("rs9841829_G", "rs29001570_T") | -0.07 | 0.51 | -0.06 (-0.18, 0.01) |
| At Least 2 Jointly Valid | -0.08 | 0.87 | NA |
| At Least 1 Marginally Valid | -0.11 | 0.88 | NA |

Supplementary table 9: Correlation between maternal and paternal genotypes in the Norwegian Mother, Father, and Child Study.

| SNP | Chromosome | MAF | HWE p-value | Batch specific imputation score (m12) | Batch specific imputation score (m24) | Batch specific imputation score (njl) | MoBa maternal-paternal correlation |
|---|---|---|---|---|---|---|---|
| rs1154447_T | 4 | 0.4521000 | 0.22620 | 0.985588 | 0.985381 | 0.792674 | -0.0005435 |
| rs11940694_A | 4 | 0.3813000 | 0.26520 | 1.000000 | 1.000000 | 1.000000 | 0.0062836 |
| rs12955142_C | 18 | 0.0462100 | 0.40140 | 0.810193 | 0.806994 | 0.976379 | 0.0070774 |
| rs140280172_A | 4 | 0.0012214 | 1.00000 | 0.715598 | 0.739144 | 0.651505 | -0.0024076 |
| rs145441283_G | 4 | 0.0018890 | 0.05071 | 0.499465 | 0.548268 | 0.530753 | -0.0040098 |
| rs145452708_C | 4 | 0.0025300 | 0.08957 | 0.752144 | 0.748261 | 0.787136 | -0.0055514 |
| rs149127347_G | 4 | 0.0009445 | 0.01268 | 0.536891 | 0.663150 | 0.657911 | -0.0022889 |
| rs29001570_C | 4 | 0.0039130 | 1.00000 | 0.710232 | 0.752623 | 0.839694 | 0.0104208 |
| rs3114045_T | 4 | 0.1210000 | 0.33430 | 0.964588 | 0.960405 | 0.969773 | 0.0078292 |
| rs9841829_G | 3 | 0.2385000 | 0.01059 | 0.999040 | 0.999213 | 0.996186 | 0.0034858 |
| rs9991733_G | 4 | 0.2709000 | 0.57460 | 0.998556 | 0.998534 | 0.958234 | 0.0035274 |

### 10 R functions for application of bounds across multiple proposed instruments

```
#load necessary packages
library(AER)
library(tidyverse)
library(xlsx)
library(writexl)
library(foreign)
library(broom)
library(nnet)
library(boot)

##additional function to get default arguments out
match.call.defaults <- function(...) {
  call <- evalq(match.call(expand.dots = FALSE), parent.frame(1))
  formals <- evalq(formals(), parent.frame(1))

  for(i in setdiff(names(formals), names(call)))
    call[i] <- list( formals[[i]] )


  match.call(sys.function(sys.parent()), call)
}

## Creating internal function to get maximum value of the instrumental
## inequalities for single given joint instrument
## NOTE: INSTRUMENT, X, and Y CAN BE MULTICATEGORICAL BUT CANNOT BE CONTINUOUS
## ARGUMENTS
## data: names of dataset (data.frame)
## instrument: name of instrument variable (character)
## x: name of exposure variable (character)
## y: name of outcome variable (character)
## weight: name of externally calculated weight variable (character)
run_instrumental_inequalities_singlejointiv <- function(data, y, x, instrument,
                                                        weight=NULL){

  # Set variable names
  data$Y <- data[[y]]
  data$X <- data[[x]]
  data$IV <- data[[instrument]]
  if(is.null(weight)){data$weight <- 1}else{data$weight <- data[[weight]]}
  n_uniq_Y <- length(unique(data$Y))

  # Creating matrix with all possible combinations of proposed IV and exposure
  com <- lapply(1:n_uniq_Y, function(k) {
    unique(data$IV)
  })
  com[[length(com)+1]] <- unique(data$X)
  com <- expand.grid(com, stringsAsFactors = FALSE)
  names(com) <- c(paste0("IV", 1:n_uniq_Y),"X")

  # Filling in com matrix with proportions from the data - values of
```

```
  # instrumental inequalities for different combinations
  prp <- lapply(1:n_uniq_Y, function(i) {
    sapply(1:nrow(com), function(j) {
      sum(data$weight[which(data$IV==com[j, i] & data$X==com$X[j] &
                             data$Y==unique(data$Y)[i])])/
        sum(data$weight[which(data$IV==com[j, i])])
    })
  }) %>% do.call(cbind, .) %>% as.data.frame
  names(prp) <- paste0("Y", unique(data$Y), "_IV", 1:n_uniq_Y)
  prp$sum_prop <- rowSums(prp)

  # Combine com matrix with proportions and values of instrumental inequalities
  com <- cbind(com, prp)
  return(com)
}


## Function applying the instrumental inequalities for a given exposure-outcome
## pair across all combinations of multiple proposed instruments.
##
## ARGUMENTS:
## datasetname: the dataset to be used (data.frame)
## IV: a character vector containing the names of the variables proposed as
## instruments (character vector)
## exposure: the name of the exposure of interest (character)
## outcome: the name of the outcome of interest (character)
## single_weight: the name of an externally calculated weight variable (character)
## count_output: options to limit additional output of function. Options are
## "unweighted" (calculates numbers of contributing rows without weights),
## "weighted" (calculates number of individuals contributing in weighted
## pseudopopulation), or "full" (gives both weighted and unweighted number of
## individuals in levels of joint instrument).
##
## The function outputs a list of 4 results - the first is a summary table of
## the findings, and the second, third, and fourth are information necessary for
## the creation of the bfi graph visualizations of the results.
## The function will remove any rows with missing data.
instrumental_inequalities_multiv <- function(datasetname, IV, exposure, outcome,
                                             input_weight = NULL,
                                             generate_weights = FALSE,
                                             weight_covs = NULL,
                                             count_output= "full"){

  k <- length(IV)

  # create list of sets of instruments
  mylist <- lapply(seq_along(IV), function(i) combn(IV, i, FUN = list))

  mylist <- flatten(mylist)

  # check GRS and add to list
  datasetname$AlleleScore <- apply(datasetname[IV], 1, sum)
  mylist <- c(mylist, "AlleleScore")
```

```r
# create summary table
summarydat <- matrix(nrow=length(mylist), ncol=11)
colnames(summarydat) <- c('Unweighted Nonzero Cell Count',
                          'Unweighted Smallest Cell',
                          'Unweighted Number Cells < 10',
                          'Weighted Nonzero Cell Count',
                          'Weighted Smallest Cell',
                          'Weighted Number Cells < 10',
                          'Bonet trichotomous inequality holds?',
                          'Balke-Pearl IV Inequalities Hold?',
                          'BP Inequalities Max Value',
                          'BP Violating Strata of Instrument',
                          'BP Violating Exposure Level')

rownames(summarydat) <- c(mylist)

#get names alone for covariates for weights generated
covnames <- weight_covs[!(str_detect(weight_covs, "\\*|\\^"))]

#create variable and run results for each possible combination of proposed IVs
for (i in 1:length(mylist)){
  dat <- datasetname %>% select(IV, everything())
  dat$a <- dat[[exposure]]
  dat$y <- dat[[outcome]]
  if(is.null(input_weight)){dat$input_weight <- 1}else{
    dat$input_weight <- dat[[input_weight]]}
  dat <- dat%>% select(IV, AlleleScore, a, y, input_weight, covnames)
  dat <- dat %>% drop_na()
  n_uniq_Y <- length(unique(dat$Y))

  #create new joint variable
  IVT = mylist[i]
  dat <- unite_(dat,"jointIV", flatten(IVT), remove = FALSE)


  #generate weights for jointIV if generated weights turned on
  if(generate_weights==TRUE && is.null(weight_covs)==FALSE &&
     length(unique(dat$jointIV)) > 2){
    denom <- reformulate(weight_covs, response = "jointIV")

    denom_obj <- multinom(denom, dat, trace=FALSE)
    p_denom <- as.data.frame(predict(denom_obj, type="probs"))
    for (g in 1:length(sort(unique(dat$jointIV)))){
      dat$denom[with(dat, jointIV == sort(unique(dat$jointIV))[g])] <-
        p_denom[ dat$jointIV == sort(unique(dat$jointIV))[g],g]
    }
    dat$gen_weight <- 1/dat$denom
  }else if(generate_weights==TRUE && is.null(weight_covs)==FALSE &&
           length(unique(dat$jointIV))<3){
    denom <- reformulate(weight_covs, response = "as.numeric(jointIV)")
    denom_obj <- glm(denom, dat, family="binomial")
    dat$denom <- predict(denom_obj, type="response")
    dat$gen_weight <- 1/dat$denom
```

```r
}else if(generate_weights==TRUE && is.null(weight_covs)==TRUE){
  dat$gen_weight <- 1
  print("No covariates supplied, IP weights set to 1")
} else{
  dat$gen_weight <- 1
}

#multiply input weight and generated weight together
dat$weight <- dat$input_weight*dat$gen_weight

#running instrumental inequalities function
combo <- run_instrumental_inequalities_singlejointiv(data=dat,
                                                     y="y", x="a",
                                                     instrument="jointIV",
                                                     weight="weight")
ineq <- aggregate(sum_prop ~ IV1, data=combo, FUN=max)$sum_prop %>% max

#print IV inequalities held or no
summarydat[i, 8] <- if (ineq<=1) {"yes"} else {"no"}
summarydat[i, 9] <- ineq

#creates dataset of violating strata
combo1 <- combo %>% filter(sum_prop > 1)

#generate list of jointIVs to be printed in violating strata
summarydat[i, 10] <- ifelse(ineq <= 1, "none",
                            paste(list(unique(flatten(flatten(
                              lapply(1:n_uniq_Y, function(j) {combo1[j]}
                                     )))))))

#number cells, smallest cells, count cells under 10 for unweighted rows
ftable <- rle(sort(dat$jointIV))
summarydat[i, 1] <- length(ftable$lengths)
summarydat[i, 2] <- min(ftable$lengths)
summarydat[i, 3] <- sum(ftable$lengths < 10)

#number cells, smallest cells, count cells under 10 with weights
#incorporated
zcount <- dat %>% group_by(jointIV) %>% tally(wt=weight)
summarydat[i, 4] <- length(zcount$n[which(zcount$n != 0)])
summarydat[i, 5] <- min(zcount$n[which(zcount$n != 0)])
summarydat[i, 6] <- sum(zcount$n < 10)

#exposure level violated
summarydat[i, 11] <- if (ineq <= 1) {"none"} else {paste(list(unique(
  combo1$X)))}

#Bonet trichotomous instrument inequality
#only eligible if binary exposure and outcome, trichotomous instrument
triineq <- ifelse(length(unique(dat$a)) > 2, "NA",
                  ifelse(length(unique(dat$y)) > 2, "NA",
                         ifelse(length(unique(dat$jointIV)) == 3,
                                sum(dat$weight[which(dat$a == min(dat$a) &
```

```
                                                dat$y == max(dat$y) &
                                                dat$jointIV ==
                                                max(as.numeric(
                                                  dat$jointIV))-1)])/
                                  sum(dat$weight[which(dat$jointIV ==
                                                  max(as.numeric(
                                                    dat$jointIV
                                                    ))-1)]) +
                                  sum(dat$weight[which(dat$a == min(dat$a) &
                                                  dat$y == min(dat$y) &
                                                  dat$jointIV == max(
                                                    as.numeric(
                                                      dat$jointIV
                                                      )))]) /
                                  sum(dat$weight[which(dat$jointIV == max(
                                    as.numeric(dat$jointIV)))]) +
                                  sum(dat$weight[which(dat$a == min(dat$a) &
                                                  dat$y ==
                                                  max(dat$y) &
                                                  dat$jointIV ==
                                                  min(as.numeric(
                                                    dat$jointIV)))])/
                                  sum(dat$weight[which(dat$jointIV ==
                                                  min(as.numeric(
                                                    dat$jointIV)))])+
                                  sum(dat$weight[which(dat$a == max(dat$a) &
                                                  dat$y ==
                                                  max(dat$y) &
                                                  dat$jointIV ==
                                                  max(
                                                    as.numeric(
                                                    dat$jointIV))-1
                                                )]) /
                                  sum(dat$weight[
                                    which(dat$jointIV == max(as.numeric(
                                      dat$jointIV))-1)]) +
                                  sum(dat$weight[which(dat$a == max(dat$a) &
                                                  dat$y ==
                                                  min(dat$y) &
                                                  dat$jointIV ==
                                                  min(as.numeric(
                                                    dat$jointIV)))])/
                                  sum(dat$weight[
                                    which(dat$jointIV == min(as.numeric(
                                      dat$jointIV)))]), "NA" )))
  summarydat[i, 7] <- ifelse(triineq<=2, "yes", triineq)
  print(paste0(i," of ",length(mylist), " combinations complete"))
}


#make copy of max values of BP inequalities
max_bp_ineqs <- unname(summarydat[, 9])
```

```
  ##options to reduce output of weighted and unweighted counts
  ##Ifelse statements allowing deletion of irrelevant columns
  summarydat <- if(count_output=="weighted"){test <- summarydat[,4:11]} else{
    if(count_output=="unweighted"){test <- summarydat[,-c(4,5,6)]} else{
      summarydat}}


  resultslist <- list(summarydat, mylist, k, max_bp_ineqs, datasetname,
                      as.list(match.call.defaults()))
  return(resultslist)
}


##Inequalities plotting function
##Arguments:
##ineq_output: output of instrumental_inequalities_multiv function
##title: title to be given to plot
##
##outputs a visualization of the application of the instrumental inequalities,
##where each row represents a proposed instrument, and each vertical line
##connects a set of variables jointly proposed as instruments. The color of each
##node represents the value of the instrumental inequalities, with white
##indicating that the instrumental inequalities held, and darker colors
##indicating increasing magnitudes of violation.

plot_instrumental_inequalities <- function(ineq_output, title){
  #set up
  instru <- ineq_output[[2]]
  k <- ineq_output[[3]]
  ineqs <- ineq_output[[4]]

  ##determining title
  if(missing(title)){title<-NA}

  ##generating nodes dataset
  nodes <- data.frame(id=1:length(flatten(instru)))
  nodes$label1 <- lapply(1:length(flatten(instru)), function(i){
    flatten(instru)[[i]]})

  ##generate y position - aligning along y spots
  nodes$y <- 1
  for (j in 1:k){nodes<-nodes %>% mutate(y=ifelse(label1==instru[[j]], j, y))}
  nodes$y <- as.numeric(nodes$y)
  nodes$y <- nodes$y*10
  ##label of group - creates their x axis coordinates
  nodes$x <- flatten(lapply(1:length(instru), function(i){rep(length(
    instru[1:i]),
                                                              length(flatten(
                                                                instru[i])))}))
  nodes$x <- unlist(nodes$x)
  nodes$x <- nodes$x*10

  ##edges generation
```

```
  edges <- data.frame(id=1:length(unique(nodes$y)))
  edges$fromy <- unique(nodes$y)
  edges$fromx <- tapply(nodes$x, nodes$y, min)
  edges$toy <- edges$fromy
  edges$tox <- tapply(nodes$x, nodes$y, max)
  vertedges <- data.frame(id=1:length(unique(nodes$x)))
  vertedges$fromx <- unique(nodes$x)
  vertedges$tox <- vertedges$fromx
  vertedges$fromy <- tapply(nodes$y, nodes$x, min)
  vertedges$toy <- tapply(nodes$y, nodes$x, max)
  edges <- rbind(edges, vertedges)

  ## size and color of nodes
  printsumnum <- function(i){print(ineqs[i])}
  nodes <- nodes %>% rowwise %>% mutate(colorfactor=printsumnum(x/10))
  nodes$ii <- ifelse(nodes$colorfactor<=1, NA, cut(as.numeric(
    nodes$colorfactor), breaks=seq(1, 2, len=100), include.lowest=TRUE))
  nodes$color <- ifelse(nodes$colorfactor<=1, colorRampPalette("grey99")(1),
                        colorRampPalette(c("gray60", "gray22"))(99)[nodes$ii])

  ##node labels
  nodes$label1 <- ifelse(nodes$id<=k, nodes$label1, ifelse(nodes$id==length(
    flatten(instru)), nodes$label1, ""))

  ##generating plot itself
  layout(matrix(1:2,nrow=1),widths=c(0.8,0.2))
  par(mar=c(5.1,4.1,4.1,1.0))
  plot(c(-2,max(nodes$x)+5),c(0,max(nodes$y)+5),type = 'n', axes = F,xlab = '',
       ylab = '', main = title)
  segments(edges$fromx, edges$fromy, x1= edges$tox, y1= edges$toy)
  points(nodes$x, nodes$y, pch=21, cex=3.5, bg=nodes$color)
  text(nodes$x[1:length(flatten(instru))-1], nodes$y[1:length(
    flatten(instru))-1],
       nodes$label1[1:length(flatten(instru))-1], pos=1, offset=1)
  text(nodes$x[length(flatten(instru))]+2, nodes$y[length(flatten(instru))],
       nodes$label1[length(flatten(instru))], pos=3, offset=1.5)
  legend_image <- as.raster(matrix(colorRampPalette(c('gray22', 'gray60'))(99),
                                   ncol=1))
  par(mar=c(5.1,1.0,4.1,2.1))
  plot(c(0,2),c(0,2),type = 'n', axes = F,xlab = '', ylab = '', main = 'Legend')
  text(x=1.5, y = c(.5, seq(1,2,by=.25)), labels = c(0, 1, seq(1.25,2,by=.25)))
  rasterImage(legend_image, 0, 1, 1,2)
  rect(0.025,.5,1,1, col='grey99', border='black')
}




###BOUNDS INCLUDING WEIGHTS
##Internal function to estimate part of bounds using same level of proposed
##k-level instrument, based on Richardson and Robins 2014
##
##ARGUMENTS:
```

```
##data: the dataset to be used (must already include values for X, Y, jointIV,
##and weight)
##x: exposure variable (binary)
##y: outcome variable (binary)
##z: a level of the single k-level proposed instrument
RRIVbounds_samez <- function(data, x,y,z){
  sum(data$weight[which(data$X == x & data$Y == y & data$jointIV == z)]) /
    sum(data$weight[which(data$jointIV == z)]) +
    sum(data$weight[which(data$X == (1-x) & data$jointIV == z)]) /
    sum(data$weight[which(data$jointIV == z)])}

##Internal function to estimate part of bounds using different levels of
##proposed k-level instrument, based on Richardson & Robins 2014
##
##ARGUMENTS:
##data: the dataset to be used (must already include values for X, Y, jointIV,
##and weight)
##x: exposure variable (binary)
##y: outcome variable (binary)
##z: a level of the single k-level proposed instrument
##s: a level of the single k-level proposed instrument that =/= z
RRIVbounds_diffz <- function(data,x,y,z,s){(sum(data$weight[
  which(data$X == x & data$Y == y & data$jointIV == z)]) /
                                          sum(data$weight[
                                            which(data$jointIV == z)])) +
    (sum(data$weight[which(data$X ==
                             (1-x) & data$Y == 0 & data$jointIV == z)]) / sum(
                               data$weight[which(data$jointIV == z)])) +
    (sum(data$weight[which(data$X == x & data$Y == y & data$jointIV == s)]) /
       sum(data$weight[which(data$jointIV == s)])) +
    (sum(data$weight[which(data$X == (1-x) & data$Y == 1 & data$jointIV == s)])/
       sum(data$weight[which(data$jointIV == s)]))}


##Function to create bounds filtered based on instrumental inequalities
##
##ARGUMENTS:
##ineq_output: output of the instrumental_inequalities_multiv function
##exclude: optional vector of proposed instruments to exclude from bounds based
##on visual inspection. Allele Score must be manually added to this vector.
##
##
##Function returns dataframe consisting of 1. possible instrument validity
##assumptions (any combinations of proposed instruments which were not
##falsified - assumed marginally and jointly valid, and sets of assumptions
##about marginal and joint validity of the set of SNPs proposed as instruments),
##2. maximum value of the instrumental inequalities for each proposed marginal
##or joint instrument, 3. lower bound on ACE from proposed instrument, and
##4. upper bound on ACE from proposed instrument.
##
##Note: While the instrumental inequalities can be applied to multicategorical
## proposed instruments, exposures, and outcomes, the Richardson-Robins bounds
##allow for multicategorical proposed instruments, but can only be used with
```

```r
##binary exposures and outcomes. If applied to a binary exposure or outcome, the
##function will return an error.
RRIVbounds <- function(ineq_output, exclude){

  ##Set up
  datasetname <- ineq_output[[5]]
  IV <- eval(ineq_output[[6]]$IV)
  x <- eval(ineq_output[[6]]$exposure)
  y <- eval(ineq_output[[6]]$outcome)
  input_weight <- eval(ineq_output[[6]]$input_weight)
  generate_weights <- eval(ineq_output[[6]]$generate_weights)
  weight_covs <- eval(ineq_output[[6]]$weight_covs)
  instru <- ineq_output[2]
  ineq <- ineq_output[4]

  ##check if exclude list contains anything
  if(missing(exclude)){exclude = c()}

  ##Combining list of possible instruments and inequalities values into a tibble
  instru <-data_frame(flatten(instru))
  ineq <- data_frame(flatten(ineq))
  bounds <- cbind(instru, ineq)
  colnames(bounds) <-c("instru", "ineq")


  ##Filter out columns where inequalities are violated
  bounds <- bounds %>% filter(ineq <= 1)

  ##Filter excluded out
  for (i in exclude){
    bounds <- bounds %>% filter(str_detect(bounds$instru,i)==FALSE)}


  ##Add columns for the bounds
  bounds <- add_column(bounds, LB=NA, UB=NA)

  datasetname$AlleleScore <- apply(datasetname[flatten(instru)[[
    length(flatten(instru))-1]]], 1, sum)

  #get names alone for covariates for weights generated
  covnames <- weight_covs[!(str_detect(weight_covs, "\\*|\\^"))]



  for (i in 1:length(bounds$instru)){
    dat <- datasetname %>% select(everything())
    dat$Y <- dat[[y]]
    if(length(unique(dat$Y))!=2){
      print("ERROR: outcome must be binary")
      break

    }
    dat$Y <- ifelse(dat$Y==min(unique(dat$Y)), 0, 1)
```

```
  dat$X <- dat[[x]]
  if(length(unique(dat$X))!=2){
    print("ERROR: exposure must be binary")
    break

  }
  dat$X <- ifelse(dat$X==min(unique(dat$X)), 0, 1)
  IVT <- bounds$instru[[i]]
  if(is.null(input_weight)){dat$input_weight <- 1}else{dat$input_weight <-
    dat[[input_weight]]}
  dat <- dat %>% select(IVT, "X", "Y", "input_weight", covnames)
  dat <- dat %>% drop_na()
  dat <- unite_(dat,"jointIV", IVT, remove = FALSE)


  #generate weights for jointIV if generated weights turned on
  if(generate_weights==TRUE && is.null(weight_covs)==FALSE && length(
    unique(dat$jointIV)) > 2){
    denom <- reformulate(weight_covs, response = "jointIV")

    denom_obj <- multinom(denom, dat, trace=FALSE)
    p_denom <- as.data.frame(predict(denom_obj, type="probs"))
    for (g in 1:length(sort(unique(dat$jointIV)))){
      dat$denom[with(dat, jointIV == sort(unique(dat$jointIV))[g])] <-
        p_denom[dat$jointIV == sort(unique(dat$jointIV))[g],g]
    }
    dat$gen_weight <- 1/dat$denom
  }else if(generate_weights==TRUE && is.null(weight_covs)==FALSE && length(
    unique(dat$jointIV))<3){
    denom <- reformulate(weight_covs, response = "as.numeric(jointIV)")
    denom_obj <- glm(denom, dat, family="binomial")
    dat$denom <- predict(denom_obj, type="response")
    dat$gen_weight <- 1/dat$denom
  }else if(generate_weights==TRUE && is.null(weight_covs)==TRUE){
    dat$gen_weight <- 1
    print("No covariates supplied, IP weights set to 1")
  } else{
    dat$gen_weight <- 1
  }

  #multiply input weight and generated weight together
  dat$weight <- dat$input_weight*dat$gen_weight



  com <- lapply(1:2, function(k) {
    unique(dat$jointIV)
  })

  com <- expand.grid(com, stringsAsFactors = FALSE)

  names(com) <- c(paste0("IV", 1:2))
  com <- com %>% filter(IV1 != IV2)
```

```
  com <- com %>% rowwise() %>% mutate( g101=RRIVbounds_samez(dat,1,0,IV1),
                                       g102=RRIVbounds_diffz(dat,1,0,IV1,IV2),
                                       g011=RRIVbounds_samez(dat, 0,1,IV1),
                                       g012=RRIVbounds_diffz(dat, 0,1,IV1,
                                                              IV2),
                                       g001=RRIVbounds_samez(dat, 0,0,IV1),
                                       g002=RRIVbounds_diffz(dat, 0,0,IV1,
                                                              IV2),
                                       g111=RRIVbounds_samez(dat, 1,1,IV1),
                                       g112=RRIVbounds_diffz(dat, 1,1,IV1,
                                                              IV2))


  bounds[i,3]<- 1- min(min(com$g102), min(com$g101))-min(min(com$g011),
                                                          min(com$g012))
  bounds[i,4] <- min(min(com$g001), min(com$g002))+min(min(com$g111),
                                                         min(com$g112))-1}

bounds <- bounds %>% mutate(inlength = lengths(instru))

bounds <- bounds %>% add_row()


for (i in 1:(max(bounds$inlength, na.rm=TRUE))) {
  bounds <- bounds %>% add_row()
}

for (i in unique(na.omit(bounds$inlength))){
  bounds$instru[length(bounds$instru)-i+1] <- sprintf("
                                                       At Least %s Jointly
                                                       Valid", i)

  bounds[length(bounds$instru)-i+1, 3] <- bounds[1:(length(na.omit(
    bounds$inlength))),] %>%
    filter(instru != "AlleleScore") %>% filter(inlength==i) %>% summarize(
      min=min(LB))
  bounds[length(bounds$instru)-i+1, 4] <- bounds[1:(length(na.omit(
    bounds$inlength))),] %>%
    filter(instru != "AlleleScore") %>% filter(inlength==i) %>% summarize(
      max=max(UB))

}

bounds$instru[length(na.omit(bounds$inlength))+1] <- "All Marginally Valid"
bounds$instru[length(bounds$instru)] <- "At Least 1 Marginally Valid"

bounds[length(na.omit(bounds$inlength))+1, 3] <- bounds[1:(length(na.omit(
  bounds$inlength))),] %>%
  filter(instru != "AlleleScore") %>% filter(inlength==1) %>% summarize(
    max=max(LB))
bounds[length(na.omit(bounds$inlength))+1, 4] <- bounds[1:(length(na.omit(
  bounds$inlength))),] %>%
```

```
    filter(instru != "AlleleScore") %>% filter(inlength==1) %>% summarize(
      min=min(UB))



  bounds <- bounds %>% select(-c(inlength))
  return(bounds)}



##Internal function to run bootstrap and get CIs for IV point estimate
##
##Arguments
##data: dataset to be used in calculation of 95% CIs for point estimate
##indices: allows boostrap to resample
##
##Function returns beta for exposure from second stage of TSLS (ACE estimate)
tsfctn <- function(data, indices){
  d <- data[indices,]
  tsls <- ivreg(Y ~ X | jointIV, data=d, weights = weight)
  return(tsls$coefficients["X"])}



##Function to add point estimates and CIs to IV bounds code filtered by
##inequalities
##
##ARGUMENTS:
##RRIVbounds_output: output of RRIVbounds function.
##ineq_output: output of the instrumental_inequalities_multiv function.
##
##Function returns dataframe consisting of 1. possible instrument validity
##assumptions (any combinations of proposed instruments which were not
##falsified - assumed marginally and jointly valid, and sets of assumptions
##about marginal and joint validity of the set of SNPs proposed as instruments),
##2. maximum value of the instrumental inequalities for each proposed marginal
##or joint instrument, 3. lower bound on ACE from proposed instrument, 4. upper
##bound on ACE from proposed instrument, 5. point estimate for ACE
##(assuming homogeneity) from proposed instrument, 6. lower 95% CI for point
##estimate for ACE (calculated using empirical bootstrap), and 7. upper
##95% CI for point estimate for ACE (calculated using empirical bootstrap).
##
##Note: Importantly, this function uses a linear-linear two stage least squares
##approach for all proposed instruments, exposures, and outcomes, including the
##all binary setting used here.
################################################################################

add_iv_est <- function(RRIVbounds_output, ineq_output){

  ##Set up
  datasetname <- ineq_output[[5]]
  IV <- eval(ineq_output[[6]]$IV)
  x <- eval(ineq_output[[6]]$exposure)
```

```r
  y <- eval(ineq_output[[6]]$outcome)
  input_weight <- eval(ineq_output[[6]]$input_weight)
  generate_weights <- eval(ineq_output[[6]]$generate_weights)
  weight_covs <- eval(ineq_output[[6]]$weight_covs)
  instru <- ineq_output[2]

##add AlleleScore to dataset again
datasetname$AlleleScore <- apply(datasetname[flatten(instru)[[length(flatten(
  instru))-1]]], 1, sum)

##add columns to combobounds results table
RRIVbounds_output <- add_column(RRIVbounds_output, est = NA, lci=NA, uci=NA)


##create list of proposed instruments that have passed inequalities
pass <- RRIVbounds_output
pass <- pass[1:length(pass$ineq[lapply(pass$ineq, length) > 0]),]
pass <- pass %>% select(instru)
pass <- pass[[1]]


#get names alone for covariates for weights generated
covnames <- weight_covs[!(str_detect(weight_covs, "\\*|\\^"))]

##for loop generating point estimates and confidence intervals for each IV in
##pass

for (i in 1:length(pass)){
  dat <- datasetname %>% select(everything())
  dat$Y <- dat[[y]]
  if(length(unique(dat$Y))!=2){
    print("ERROR: outcome must be binary")
    break

  }
  dat$Y <- ifelse(dat$Y==min(unique(dat$Y)), 0, 1)
  dat$X <- dat[[x]]
  if(length(unique(dat$X))!=2){
    print("ERROR: outcome must be binary")
    break

  }
  dat$X <- ifelse(dat$X==min(unique(dat$X)), 0, 1)
  IVT <- pass[[i]]
  if(is.null(input_weight)){dat$input_weight <- 1}else{dat$input_weight <- dat[[
    input_weight]]}
  dat <- dat %>% select(IVT, "X", "Y", "input_weight", covnames)
  dat <- dat %>% drop_na()
  dat <- unite_(dat,"jointIV", IVT, remove = FALSE)


  #generate weights for jointIV if generated weights turned on
  if(generate_weights==TRUE && is.null(weight_covs)==FALSE && length(unique(
```

```
    dat$jointIV)) > 2){
    denom <- reformulate(weight_covs, response = "jointIV")

    denom_obj <- multinom(denom, dat, trace=FALSE)
    p_denom <- as.data.frame(predict(denom_obj, type="probs"))
    for (g in 1:length(sort(unique(dat$jointIV)))){
      dat$denom[with(dat, jointIV == sort(unique(dat$jointIV))[g])] <-
        p_denom[dat$jointIV == sort(unique(dat$jointIV))[g],g]
    }
    dat$gen_weight <- 1/dat$denom
  }else if(generate_weights==TRUE && is.null(weight_covs)==FALSE && length(
    unique(dat$jointIV))<3){
    denom <- reformulate(weight_covs, response = "as.numeric(jointIV)")
    denom_obj <- glm(denom, dat, family="binomial")
    dat$denom <- predict(denom_obj, type="response")
    dat$gen_weight <- 1/dat$denom
  }else if(generate_weights==TRUE && is.null(weight_covs)==TRUE){
    dat$gen_weight <- 1
    print("No covariates supplied, IP weights set to 1")
  } else{
    dat$gen_weight <- 1
  }

  #multiply input weight and generated weight together
  dat$weight <- dat$input_weight*dat$gen_weight


  #run IV estimation (2 stage least squares)
  tsls <- ivreg(Y ~ X | jointIV, data=dat, weights = weight)
  RRIVbounds_output$est[i] <- tsls$coefficients["X"]

  bootres <- boot(data=dat, statistic = tsfctn, R=1000)
  bootcis <- boot.ci(bootres, type="basic")
  RRIVbounds_output$lci[i] <- bootcis$basic[4]
  RRIVbounds_output$uci[i] <- bootcis$basic[5]}

return(RRIVbounds_output)}

##load necessary dataset

load(file="./data/genlinked_20200131.RData")

##running functions on data
##using rs12955142 as proxy from chr18:72124965
#instrumental inequalities
exposurelist <- c("any_alc", "mod_alc")
outcomelist <- c("adh98")
covlist <- list(NULL, c("PC1", "PC2", "PC3", "PC4", "PC5", "PC6", "PC7", "PC8",
                        "PC9", "PC10"))
expout <- list(exposurelist, outcomelist, covlist)
expout <- expand.grid(expout, stringsAsFactors = FALSE)

genlist <- c(FALSE, FALSE, TRUE, TRUE)
```

```r
expout <- cbind(expout, genlist)

names(expout) <- c("exposure", "outcome", "covs", "genweight")


plotnames <- c( "output/anyalc_adh98_nopc_bounds.png",
                "output/modalc_adh98_nopc_bounds.png",
                "output/anyalc_adh98_pc_bounds.png",
                "output/anyalc_adh98_pc_bounds.png")


#remove missings on variables other than exposure

newdata <- newdata %>% select("PREG_ID_2306", "IID", "any_alc", "mod_alc",
                              "adh98", "rs11940694_A", "rs9991733_G",
                              "rs145452708_C", "rs3114045_T", "rs1154447_T",
                              "rs9841829_G", "rs145441283_G", "rs29001570_C",
                              "rs140280172_A", "rs149127347_G", "rs12955142_C",
                              "PC1", "PC2", "PC3", "PC4", "PC5", "PC6", "PC7",
                              "PC8", "PC9", "PC10")





newdata <- newdata %>% drop_na("PREG_ID_2306", "IID", "any_alc", "adh98",
                               "rs11940694_A", "rs9991733_G", "rs145452708_C",
                               "rs3114045_T", "rs1154447_T", "rs9841829_G",
                               "rs145441283_G", "rs29001570_C", "rs140280172_A",
                               "rs149127347_G", "rs12955142_C", "PC1", "PC2",
                               "PC3", "PC4", "PC5", "PC6", "PC7", "PC8", "PC9",
                               "PC10")

for (i in 1:nrow(expout)){
  #drop missing data on exposure
  newdata2 <- newdata %>% drop_na(expout$exposure[i])

  #run inequalities
  ineq_res <- instrumental_inequalities_multiv(datasetname = newdata2, IV=
                                                 c("rs11940694_A",
                                                   "rs9991733_G",
                                                   "rs145452708_C",
                                                   "rs3114045_T",
                                                   "rs1154447_T", "rs9841829_G",
                                                   "rs145441283_G",
                                                   "rs29001570_C",
                                                   "rs140280172_A",
                                                   "rs149127347_G",
                                                   "rs12955142_C"), exposure =
                                                 expout$exposure[i],
                                               outcome = expout$outcome[i],
                                               input_weight = NULL,
                                               generate_weights =
```

```
                                                      expout$genweight[i],
                                                    weight_covs = expout$covs[i],
                                                    count_output = "full")

  #plot and save instrumental inequalities

  eval(parse(text=sprintf("png(
  'output/%s_%s_%s_pc%s_instrumentalinequalities.png', width=30000, height=700)
                        plot_instrumental_inequalities(ineq_output = ineq_any,
                        title='Instrumental Inequalities for MR model for
                        effect of %s on %s')
                        dev.off()", Sys.Date(), expout$exposure[i],
                        expout$outcome[i], expout$genweight[i],
                        expout$exposure[i], expout$outcome[i])))


  write_xlsx(ineq_res, path = paste0('output/', Sys.Date(), expout$exposure[i],
                                    expout$outcome[i], "pc",
                                    expout$genweight[i] ,"_ineq.xlsx"))


  ##IV bounds
  bounds_res<- RRIVbounds(ineq_res, exclude = NULL)

  ##add iv estimates
  bounds_points <- add_iv_est(bounds_res, ineq_res)

  ##plot bounds
  png(plotnames[i], width = 1385, height = 750)
  plot(0,0,type='n',xlim=c(-1.3,1),ylim=c(21,0),ylab='',
       xlab="Average causal effect of any vs. no alcohol consumption in
       pregnancy on offspring ADHD", yaxt='n')
  abline(v=0,lty=1)
  for (j in 1:length(bounds_points$instru)){
    text(-1.1, j, labels = bounds_points$instru[j], cex=.5)
    segments(x0 = bounds_points$LB[j], y0 = j, x1 = bounds_points$UB[j])
    segments(x0 = bounds_points$lci[j], y0 = j+0.5, x1 = bounds_points$uci[j],
             lty = "dashed", lwd = 1)
    points(x = bounds_points$est[j], y = j+0.5, pch=19)}
  dev.off()


  ##save bounds points file
  write_xlsx(bounds_pts, path = paste('output/', Sys.Date(),
                                     expout$exposure[i],
                                     expout$outcome[i], "pc",
                                     expout$genweight[i],
                                     "_boundspts.xlsx", sep = "_"))

}


####RUNNING INEQUALITIES WITH MULTICATEGORICAL EXPOSURES
```

```r
exposurelist <- c("alc3cat", "alc4cat", "alc7cat")
outcomelist <- c("adh98")
covlist <- list(NULL, c("PC1", "PC2", "PC3", "PC4", "PC5", "PC6", "PC7", "PC8",
                        "PC9", "PC10"))
expout <- list(exposurelist, outcomelist, covlist)
expout <- expand.grid(expout, stringsAsFactors = FALSE)

genlist <- c(FALSE, FALSE, TRUE, TRUE)
expout <- cbind(expout, genlist)

names(expout) <- c("exposure", "outcome", "covs", "genweight")



#remove missings on variables other than exposure

newdata <- newdata %>% select("PREG_ID_2306", "IID", "alc3cat", "alc4cat",
                              "alc7cat", "adh98",
                              "rs11940694_A", "rs9991733_G", "rs145452708_C",
                              "rs3114045_T", "rs1154447_T", "rs9841829_G",
                              "rs145441283_G", "rs29001570_C", "rs140280172_A",
                              "rs149127347_G", "rs12955142_C", "PC1", "PC2",
                              "PC3", "PC4", "PC5", "PC6", "PC7", "PC8", "PC9",
                              "PC10")





newdata <- newdata %>% drop_na("PREG_ID_2306", "IID", "alc3cat", "alc4cat",
                               "alc7cat", "adh98",
                               "rs11940694_A", "rs9991733_G", "rs145452708_C",
                               "rs3114045_T", "rs1154447_T", "rs9841829_G",
                               "rs145441283_G", "rs29001570_C", "rs140280172_A",
                               "rs149127347_G", "rs12955142_C", "PC1", "PC2",
                               "PC3", "PC4", "PC5", "PC6", "PC7", "PC8", "PC9",
                               "PC10")

for (i in 1:nrow(expout)){
  #drop missing data on exposure
  newdata2 <- newdata %>% drop_na(expout$exposure[i])

  #run inequalities
  ineq_res <- instrumental_inequalities_multiv(datasetname = newdata2, IV=
                                                 c("rs11940694_A",
                                                   "rs9991733_G",
                                                   "rs145452708_C",
                                                   "rs3114045_T",
                                                   "rs1154447_T",
                                                   "rs9841829_G",
                                                   "rs145441283_G",
                                                   "rs29001570_C",
                                                   "rs140280172_A",
```

```
                                                "rs149127347_G",
                                                "rs12955142_C"), exposure =
                                              expout$exposure[i],
                                            outcome = expout$outcome[i],
                                            input_weight = NULL,
                                            generate_weights =
                                              expout$genweight[i],
                                            weight_covs = expout$covs[i],
                                            count_output = "full")

  #plot and save instrumental inequalities

  eval(parse(text=sprintf("png(
  'output/%s_%s_%s_pc%s_instrumentalinequalities.png', width=30000, height=700)
                          plot_instrumental_inequalities(ineq_output = ineq_any,
                          title='Instrumental Inequalities for MR model
                          for effect of %s on %s')
                          dev.off()", Sys.Date(), expout$exposure[i],
                          expout$outcome[i], expout$genweight[i],
                          expout$exposure[i], expout$outcome[i])))


  write_xlsx(ineq_res, path = paste0('output/', Sys.Date(), expout$exposure[i],
                                     expout$outcome[i], "pc",
                                     expout$genweight[i] ,"_ineq.xlsx"))
}
```

Faassen, Ernst van, and Onni Niemelä. 2011. *Biochemistry of Prenatal Alcohol Exposure*. New York: Nova Biomedical Books.

Helgeland, Øyvind, Marc Vaudel, Petur B Juliusson, Oddgeir Lingaas Holmen, Julius Juodakis, Jonas Bacelis, Bo Jacobsson, et al. 2019. "Genome-Wide Association Study Reveals Dynamic Role of Genetic Variation in Infant and Early Childhood Growth." *Nature Communications* 10 (1): 1–10.

Hernan, Miguel A, and James M Robins. 2020. *Causal Inference:What If*. Boca Raton: Chapman & Hall/CRC.

Johnston, KM, P Gustafson, AR Levy, and P Grootendorst. 2008. "Use of Instrumental Variables in the Analysis of Generalized Linear Models in the Presence of Unmeasured Confounding with Applications to Epidemiological Research." *Statistics in Medicine* 27 (9): 1539–56.

Kleiber, Christian, Achim Zeileis, and Maintainer Achim Zeileis. 2020. "Package 'Aer'." *R Package Version 1.2* 4.

Pierce, Brandon L, and Tyler J VanderWeele. 2012. "The Effect of Non-Differential Measurement Error on Bias, Precision and Power in Mendelian Randomization Studies." *International Journal of Epidemiology* 41

(5): 1383–93.

Rassen, Jeremy A, Sebastian Schneeweiss, Robert J Glynn, Murray A Mittleman, and M Alan Brookhart. 2009. "Instrumental Variable Analysis for Estimation of Treatment Effects with Dichotomous Outcomes." *American Journal of Epidemiology* 169 (3): 273–84.

Richardson, Thomas S., and James M. Robins. 2014. "ACE Bounds; Sems with Equilibrium Conditions." *Statistical Science* 29 (3): 363–66.

Ripley, Maintainer Brian. 2010. "Package 'Boot'." *Beaver* 9.

Robins, James M. 1997. "Marginal Structural Models." *Proceedings of the Section on Bayesian Statistical Science*, 1–10.

Swanson, Sonja A, Miguel A Hernán, Matthew Miller, James M Robins, and Thomas S Richardson. 2018. "Partial Identification of the Average Treatment Effect Using Instrumental Variables: Review of Methods for Binary Instruments, Treatments, and Outcomes." *Journal of the American Statistical Association* 113 (522): 933–47.

Wang, Linbo, and Eric Tchetgen Tchetgen. 2018. "Bounded, Efficient and Multiply Robust Estimation of Average Treatment Effects Using Instrumental Variables." *Journal of the Royal Statistical Society. Series B, Statistical Methodology* 80 (3): 531.
